# GHOSTS: Validated generation of synthetic hospital time series

**DOI:** 10.1101/2024.10.29.24316332

**Authors:** Rustam Zhumagambetov, Niklas Giesa, Sebastian D. Boie, Stefan Haufe

**Affiliations:** Mathematical Modelling and Data Analysis Department, Physikalisch-Technische Bundesanstalt (PTB), Abbestraße 2-12, Berlin, 10587, Germany; Institute of Medical Informatics, Charité – Universitätsmedizin Berlin, Invalidenstraße 90, Berlin, 10117, Germany; Uncertainty, Inverse Modeling and Machine Learning Group, Technische Universität Berlin, Marchstr 23, Berlin, 10587, Germany

**Author notes:** Corresponding author *Email address:* (Rustam Zhumagambetov).

**Keywords:** generative models, generative adversarial model, electronic health records, synthetic data, time series, privacy

## Abstract

Machine learning (ML) holds great promise to support, improve, and automatize clinical decision-making in hospitals. Model training on abundantly available routine data, however, is hindered by data protection regulations. Generative models can comply with privacy laws by learning to synthesize hospital data from a target population while ensuring data privacy. Clinical time series acquired during intensive care are, however, difficult to model using established techniques, especially due to uneven sampling intervals. Here, we introduce GHOSTS (Generator of Hospital Time Series), a novel method that is capable of generating realistic heterogeneous patient trajectories in hospitals including time series with uneven sampling intervals and static patient attributes. To achieve this, GHOSTS introduces novel regularizers and a postprocessing module leveraging low-dimensional summary statistics. We further present a suite of novel benchmarks for synthetic hospital time series, GHOSTS-Bench. We train GHOSTS on a large cohort of patient data from the MIMIC-IV and eICU critical care datasets. Along with measuring the quality of the generated data in terms of how faithfully the distributions of the real data as well as their spatio-temporal dynamics are preserved, we also measure how well ML models trained on the generated data can solve a clinical prediction task on the real data. We observe that GHOSTS outperforms two state-of-the-art approaches, DoppelGANger and HALO, with respect to these criteria. We make the GHOSTS model, a corpus of synthetic data as well as Python codes implementing GHOSTS and GHOSTS-Bench publicly available. These resources will become instrumental in the future development of powerful predictive models for intensive and perioperative care.

## 1. Introduction

Intensive care units (ICUs) are among the most highly digitalized wards in hospitals, continuously aggregating information about a patient’s vital signs, laboratory measurements, treatments, and diagnostic labels, among other data. Combined with demographic information and patient data, these data form a patient’s electronic health record (EHR). Modern machine learning (ML) models trained on routinely collected EHR data have recently achieved remarkable success in predicting severe outcomes such as acute kidney injury (Tomašev et al., 2019), different complications during perioperative care (Meyer et al., 2018; Giesa et al., 2024a,b), and death (Lichtner et al., 2021; Nistal-Nuño, 2022).

Sharing medical data across institutions is critical for the development of powerful ML models. This typically requires that the data are sufficiently “de-identified”, which is often a non-trivial task. A promising approach to data sharing is the creation of synthetic data that closely follow the distributions of the intended target population while at the same time preventing deidentification. Synthetic data can be generated either in a model-driven or a data-driven way. Model-driven approaches include physical or computational simulation. By building a physical model of human organs it becomes to a certain extent possible to imitate basic functions of the body to obtain realistic surrogates of physiological measurements (Maglio et al., 2021). However, while for certain bodily functions, organs, and systems, detailed biophysical models exist (Maglio et al., 2021), no single biophysical model can describe the complete data that is collected from patients in intensive care units (ICU). On the other hand, generative ML models are able to learn complex distributions from training data. Given a trained generative model, one can easily sample large amounts of synthetic data from that distribution.

For powerful generative models, there is a possibility that generated samples too closely resemble samples from the training data, raising potential privacy concerns similar to those arising when sharing original data. To comply with privacy laws like GDPR (General Data Protection Regulation, European Parliament and Council of the European Union, 2016) or HIPAA (Health Insurance Portability and Accountability Act, Centers for Medicare & Medicaid Services, 1996), several strategies exist. First, informed consent can be obtained from all patients in the training dataset. This is often not possible in a critical care setting, though. An alternative would be to train the model on already anonymized data, such as data that has been k-anonymized (Samarati and Sweeney, 1998). Due to the concomitant loss of information, the expressiveness of such as a model may, however, be compromised. Finally, it may be possible to show that the generated synthetic samples cannot be linked back to the individuals in the training set using membership inference attacks (MIA) (Niu et al., 2024). In the latter two cases, the synthetic data could be considered anonymized and would not fall under either of the privacy regulations.

Generative models have demonstrated excellent performance in the image processing domain. However, only few models have been proposed to synthesize EHR data collected in ICUs (Yale et al., 2020; Lin et al., 2020a; Li et al., 2023; Yoon et al., 2023). A potential reason is the heterogeneity of ICU data, which is difficult to model. While some of the collected variables are static (such as demographic and diagnostic labels), the majority of vital signs, laboratory values, and procedures/medications are assessed continuously with distinct mutually inconsistent uneven sampling rates. Another distinction is between continuous-valued features such as blood pressure on one hand and discrete features such as ICD (International Statistical Classification of Diseases and Related Health Problems, World Health Organization (WHO), 2022) codes for clinical diagnoses and procedures. A generative model that is currently capable of jointly synthesizing continuous-valued and discrete static and time series data is the DoppelGANger model (Lin et al., 2020a). However, being designed for the generation of longitudinal networking data, DoppelGANger does not have a built-in mechanism to faithfully reproduce the temporal and spectral characteristics of ICU time series, which includes artifacts introduced by uneven sampling.

### 1.1. Requirements on synthetic hospital data

In order to aid the development of ML models for clinical prediction tasks, synthetic data should fulfill various requirements (Lin et al., 2020a). Here, we focus on the following *key quality aspects*.

#### Faithfulness

The generated data should match the distribution of the training data.

#### Diversity

Generated data should reflect the full range of patients in the studied cohort, that is, faithfully cover the dispersion and the possible presence of multiple modes and clusters of the training data distribution. While being implied by faithfulness, this property is often violated in practice, where generated data are spread too tightly around a few cluster centers or even individual training samples (mode collapse).

#### Utility for downstream prediction tasks

discriminative predictive models trained on the synthetic data should perform well on corresponding real data. While this is again implied by faithfulness, it emphasizes that clinically relevant differences in the data, which could be subtle in relation to the overall variability in the training data, should be preserved.

#### Privacy

One central aspect of privacy in synthetic data generation is resilience to membership inference attacks. A MIA aims to determine if a certain data point was used to train the model. Such information could be used by various types of attackers in various ways. For instance, an attacker may use it to identify sensitive attributes: a successful MIA applied a dataset of cancer patients will reveal that the identified patient was diagnosed with cancer. Conversely, oversight bodies may use it to test if the data of a non-consenting patient has been used to train a model.

Whether generated data meet these requirements can be quantitatively assessed using appropriate metrics. However, there are currently no established procedures to measure the quality specifically of synthetic ICU time series data along the above dimensions. Addressing these research gaps, the present work makes the following *contributions*.

1. We propose a novel generative model architecture, Generator of Hospital Time Series (GHOSTS), that can generate realistic heterogeneous EHR data consisting of unevenly sampled time series and static attributes that can be either categorical, discrete, or continuous-valued. We train GHOSTS on two large cohorts of patients, the public MIMIC IV (Johnson et al., 2023) and eICU (Pollard et al., 2018) databases.
2. We devise experiments and quantitative metrics to assess the quality of generative models of EHR time series data with respect to faithfulness, diversity, utility, and privacy. We use these metrics to benchmark the synthetic MIMIC and eICU data generated by GHOSTS in comparison to data generated by the DoppelGANger and HALO methods.
3. We make the GHOSTS and GHOSTS-Bench source codes, the MIMIC-GHOSTS model, and the MIMIC-GHOSTS-DB dataset publicly available.

The remainder of the paper is structured as follows. In Section 2 (Methods), we discuss existing approaches for generating synthetic EHR data and define a set of quality dimensions and desiderata for synthetic EHR data. We also present the technical details of the GHOSTS model, the procedure to train GHOSTS to generate synthetic MIMIC data, and the quantitative metrics to measure faithfulness, diversity, utility, and privacy. In Section 3 (Results), we present the experimental details and results of a comprehensive evaluation of the GHOSTS, DoppelGANger, and HALO models. The paper ends with a discussion of the results and concluding remarks in Section 4 (Discussion) and Section 5 (Conclusion).

### 2 Methods

#### 2.1. Training data

##### MIMIC-IV

MIMIC IV (Johnson et al., 2023) is a large database containing deidentified data of more than 300,000 hospital patients (Johnson et al., 2023). To extract EHR data from the MIMIC IV tables, we used routines from the MIMIC IV companion repository (Johnson et al., 2018, 2024). We removed outliers by only keeping values within physiological ranges as defined in Harutyunyan et al. (2019). We extracted the following |*F*| = 9 ICU time series features: *F* = {SOFA score, diastolic blood pressure, systolic blood pressure, heart rate, respiratory rate, SpO2, temperature, sodium, and glucose}. These features were selected due to their high importance for clinical decision making in ICUs (Ferreira et al., 2001; Yee, 2023). Basic statistics can be found in Table S1. An ICU stay is defined as the time between the first and last available measurement from the heart rate monitor. We extracted signals within the first 48 hours of the ICU stay. We excluded stays that had any of the ICU time series features completely missing or a duration of less than 48 hours, leaving 46,337 ICU stays of 35,794 patients. In addition, we also extracted static patient attributes including patient age and gender, and the sequential organ failure assessment (SOFA) score (Vincent et al., 1996), used for estimating patient’s illness severity. The prediction of SOFA scores defines the downstream task we use to measure the utility of the generated synthetic data. The SOFA score was extracted using SQL queries provided as part of MIMIC IV (Johnson et al., 2023). The score was evaluated hourly between 24 and 48 hours after ICU admission using data of the preceding 24-hour window. To prepare the data in a suitable format for training ML models, it is necessary to harmonize the different irregular sampling rates of the individual measurements. By choosing a small-enough fixed sampling grid, uneven sampling is reflected by stepwise constant behavior of the resulting time series, where steps can occur in irregular intervals. The 48 hours worth of data were upsampled by assigning each 10-min interval the value of the closest temporally preceding measurement, and interpolated to a regular sampling interval of 10 minutes using forward fill. This resulted in |*T*| = 288 time points; where *T* = {*t*_*i*_} denotes the sequence of time points. Next, we applied forward fill followed by median imputation on the holdout set of 1000 patients (1283 ICU stays) to fill in missing values, leaving 34,641 patients.

We split data into training and testing folds based on the patients’ identifiers. We extracted *N*_train_ = 21, 057 and *N*_val_ = 1, 282 ICU stays from 16,320 and 1,000 patients for the training and validation splits, respectively, as well as *N*_test_ = 22, 463 disjoint stays from 17,321 patients for the testing split. Each ICU stay comprises of a |*F*| × |*T*| matrix **d**_time_ = (*d*_*f,t*_) of time series data, where *f* ∈ *F* indexes feature and *t* ∈ *T* is a time index. In addition, each ICU stay is associated with static patient attributes **d**_attr_ = (*g, a*), where *g* ∈ {*F, M*} is biological sex, and *a* ∈ ℕ^+^ is the patient age at the time of ICU admission. The MIMIC training and test datasets are denoted by 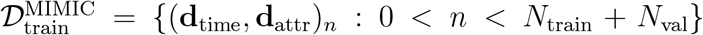 and 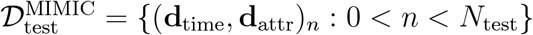, where 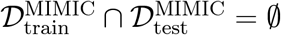.

###### eICU

eICU is a large database containing data from intensive care units across several medical centers in the United States (Pollard et al., 2018). To extract data from eICU, we used the R package ricu (Bennett et al., 2023). We used the same extraction parameters as for the MIMIC-IV database – at least 48 hours long ICU stays upsampled to 10 min intervals. We extracted 8353 stays from 8163 patients that satisfied our criteria. Then, we removed outliers, extracted features, and imputed missing values in analogy to procedures applied to the MIMIC-IV data. This resulted in 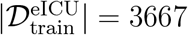, 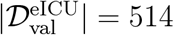, and 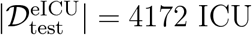 stays.

#### 2.2. Generative modeling of EHR time series

The generation of synthetic data is an unsupervised problem in which a model learns the probability distribution of a set of interdependent variables from observed training data. Suppose we have training data 𝒟^train^ = {**d**_*i*_ : **d**_*i*_ ∈ 𝔻 ⊆ ℝ^*R*^, 0 < *i* < *N*_train_, *R* ∈ ℕ^+^} sampled from a distribution *P*_true_. To empirically estimate *P*_true_, we define a parametric family of probability densities, 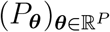, (Arjovsky et al., 2017). The task is then to find the parameter vector ***θ*** that maximizes the log-likelihood of the training data, that is,

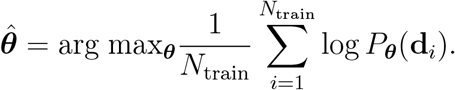

Generative ML models implicitly represent *P*_***θ***_ by a learnable generator function *G*_***θ***_ : ℤ → 𝔻 that maps random inputs **z** ∈ ℤ ⊆ ℝ^*Q*^ to samples 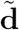.

#### 2.3. Prior work

Most state-of-the art methods for EHR generation are based on generative ML modeling, with the exception of Synthea (Walonoski et al., 2018).

##### Synthea

Synthea (Walonoski et al., 2018) is a framework for the generation of “synthetic patients”. Synthea collects summary statistics of real-world patients, such as means and percentiles, and combines them with hand-crafted rules encoded in state machines to generate synthetic static patient data. By using only aggregate summary statistics, it aims to be private by construction. This approach ensures that the generated data are somewhat consistent with the underlying data distribution at the level of individual features. However, complex distributions, involving, for example, multiple modes or higherorder interactions between features are not adequately modeled, limiting the realism of the data generated by Synthea. In particular, the generating of non-prototypical patient trajectories requires non-trivial effort, thus limiting Synthea’s usability.

##### medGAN

MedGAN (Choi et al., 2017) is a GAN-based approach that uses a combination of a GAN and an autoencoder to generate discrete features such as diagnosis or medication/treatment codes. medGAN can also generate timing information for such features but is unable to generate complete time series for continuous-valued quantities (e.g., vital signs), limiting its applicability to generate heterogeneous ICU data. The autoencoder network in medGAN is fitted on the discrete training data by minimizing the reconstruction loss. The decoder part is then applied to the generator’s output to discretize it. Finally, the discriminator differentiates the *real* input and the discrete output. The data are expressed as fixed-length vectors 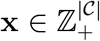, where 𝒞 is a set of features, and the value of each individual dimension indicates a count, e.g. age in years. Binary features (e.g., sex) are represented as **x** ∈ {0, 1}^|𝒞|^.

##### HealthGAN

An improvement upon medGAN is HealthGAN (Yale et al., 2020). It uses the state-of-the-art WGAN architecture (Gulrajani et al., 2017) to allow the generation of continuous and discrete data. However, as both the generator and the discriminator consist of multi-layer perceptrons with fully-connected layers, HealthGAN is not suitable to model longitudinal time-series data.

##### EHR-M-GAN

EHR-M-GAN (Li et al., 2023) is able to generate both continuous- and discrete-valued longitudinal data. Just as MedGAN, it combines the GAN approach with a variational autoencoder while also using self-supervised learning to extract common representations from correlated data of mixed-types. In addition, the generator of the GAN uses a bilateral LSTM to accommodate additional information provided by the autoencoder. The limitation of EHR-M-GAN is, however, that is *unable to generate static attributes* like sex and age.

##### HALO

The Hierarchical Autoregressive Language Model (HALO) (Theodorou et al., 2023) is a transformer model proposed for the generation of longitudinal, high-dimensional EHR time series data. It combines a language model with an autoregressive linear model to train on tokens of discrete and discretized continuous values. Tokens are combined in visits that reflect diagnoses, procedures, and medications. The model is trained in an autoregressive fashion.

##### EHR-SAFE

EHR-Safe (Yoon et al., 2023) is a generative modeling framework for generating synthetic EHR data. It combines two models: a sequential encoder-decoder network and a generative adversarial network. It can generate heterogeneous, sparse, and time-varying features with varying sequence lengths. To achieve this, a sequential encoder-decoder network is trained to learn projection of EHR data into a low-dimensional embedding space.

##### GANs

Established families of generative models include generative adversarial network (GAN, Goodfellow et al., 2014), variational autoencoders (Kingma and Welling, 2019), and diffusion-based models (Sohl-Dickstein et al., 2015). In the context of EHR generation, predominantly GANs have been used (Choi et al., 2017; Yale et al., 2020; Jarrett et al., 2021; Lu et al., 2024; Li et al., 2023; Yoon et al., 2023,, see above). A GAN consists of two parts: the generator *G*_***θ***_ and a discriminator *D*_***ϑ***_, where ***ϑ*** ∈ ℝ^*S*^. The discriminator is a classifier that tries to distinguish generated from real training samples, thereby evaluating the realism of the generated data. The aim of *G*_***θ***_ is then to minimize the loss of *D*_***ϑ***_, and the objective of *D*_***ϑ***_ is to maximize the loss of *G*_***θ***_. The two networks are, thus, competing with each other, as reflected by the min-max GAN cost function

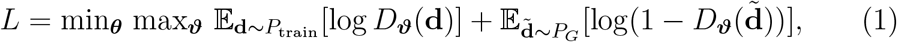

where *P*_train_ is the empirical training data distribution, *D*_***ϑ***_(**d**) is the estimated probability that **d** is a sample from *P*_train_, *P*_*G*_ is the distribution obtained by sampling the generator via 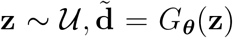, 𝒰 is a uniform distribution, and 𝔼[·] denotes expectation.

It is well-known that GANs can suffer from mode collapse and the vanishing gradient problem (e.g., Arjovsky et al., 2017). To overcome these problems, improvements to the original GAN model were suggested (Arjovsky et al., 2017; Gulrajani et al., 2017). Arjovsky et al. (2017) introduced the Wasserstein GAN (WGAN), demonstrating that the use of the Wasserstein distance as a loss function can mitigate mode collapse. Unfortunately, precise computation of Wasserstein distances is intractable. Instead, the Kantorovich-Rubinstein duality, which holds for K-Lipschitz functions, is used to approximate it (Arjovsky et al., 2017). The resulting loss minimized by WGANs is

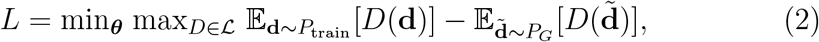

where ℒ is a set of 1-Lipschitz functions. Practically, replacing the GAN by the WGAN loss function implies that the discriminator is not anymore required to output strict probabilities between 0 to 1. This is considered to prevent both the vanishing gradients and mode collapse problems (Arjovsky et al., 2017). However, the loss formulation preserves its guarantee only under the assumption that the discriminator is K-Lipschitz. To ensure this, Arjovsky et al. (2017) proposed to clamp the discriminator weights after each update. Further work by Gulrajani et al. (2017) implements weight clipping by introducing an additional penalty on the gradient norm for random samples from the training distribution, leading to the extended loss function

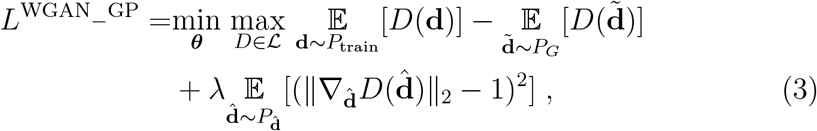

where *λ* is a scaling coefficient and 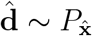 is a point uniformly sampled to lie between a random pair of points drawn from *P*_train_ and *P*_*G*_.

GAN architectures have been introduced and extensively used for image generation tasks but the concepts have also been extended to EHR data. Most existing approaches are, however, restricted to generating either static EHR attributes or time series data but not both. We discuss these in Section 2.3. DoppelGANger (DG, Lin et al., 2020a) is a WGAN with an additional gradient penalty (Gulrajani et al., 2017) designed for the generation of longitudinal networking data. Its loss is, therefore, equivalent to Equation (3). DG decouples the generation of static and time series data by first generating synthetic static data, then conditioning the time series generation on the newly generated static data. To achieve that, it uses dense layers for the generation of the static features (tabular data) and a long short-term memory (LSTM) network (Hochreiter and Schmidhuber, 1997) for the generation of the time series. Second, there are two discriminators: one for the combined static and time series data, and another one only for the static attributes.

#### 2.4. Proposed Method: Generator of Hospital Time Series (GHOSTS)

GHOSTS is an extension of the DG method designed to overcome its limitations when applied to ICU time series. To ensure the conservation of characteristic spatio-temporal dynamics in the generated time series, GHOSTS employs additional mechanisms. Most importantly, we add up to three regularizing terms to the original DG loss function. The GHOSTS cost function reads

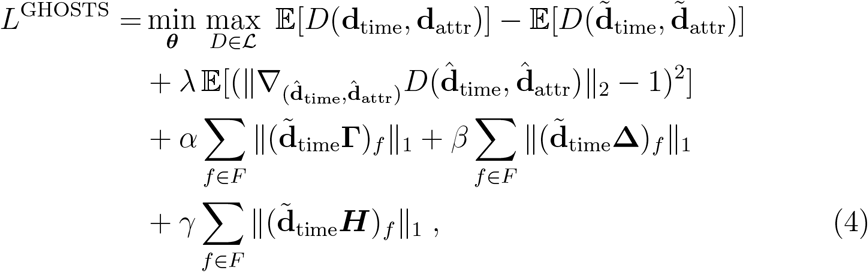

where (**d**_time_, **d**_attr_) ∼ *P*_train_, 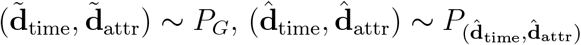, *α, β*, and *γ* are positive scaling coefficients, and ∥ · ∥_1_ denotes the *ℓ*_1_-vector-norm. **Γ** is the |*T* | × |*T* | linear type-II discrete cosine transform (DCT) matrix (Ahmed et al., 1974) with entries 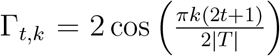. Similarly, **Δ** denotes the |*T* | × (|*T* | − 1) discrete first-order differencing operator with respect to the time dimension with entries Δ_*t,k*_ = −1 if *t* = *k*, Δ_*t,k*_ = 1 if *t* = *k* + 1, and Δ_*t,k*_ = 0 otherwise. Finally, the |*T* | × |*T* | Haar transform matrix *H* (Strang, 199*3) contains ψ*_*j,k*_(*x*) entries for 0 ≤ *j* ≤ log(|*T* |), where *j* ∈ *N* ^0^, 0 ≤ *l* ≤ 2^*j*^ − 1 and *ψ*_*j,k*_ (*x*) = *ψ*(2^*j*^*x* − *k*), 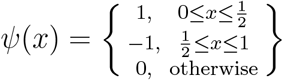. **The** last three terms of (4) are novel regularizers enforcing stepwise behavior, thereby modeling uneven sampling intervals. The fourth term encourages sparsity of each individual time series feature in the spectral domain. This is achieved by penalizing the *ℓ*_1_-norm of the coefficients of a temporal discrete cosine transform, which drives some frequency coefficients to zero. While the spectrum is not supposed to be completely sparse, it is expected to be dominated by higher harmonic frequencies of each feature’s individual sampling frequency. Frequencies in between these higher harmonics are, therefore, expected to be absent, implying the respective DCT coefficients to be zero. The fifth term penalizes the *ℓ*_1_-norm of the 1^st^ temporal derivative of each time series. This leads to sparsification of first-order differences meaning that, with *β* chosen to be appropriately high, most temporal differences of the generated data will be zero (c.f. the concept of total variation denoising, Rudin et al., 1992). In the time domain, this corresponds to piecewiese constant time courses, which is consistent with the structure of the data (c.f., Figure 1). This penalty, thus, encodes the preferences for sampling intervals larger than 10 minutes while allowing for uneven and inconsistent sampling schemes across features. The sixth term penalizes the *ℓ*_1_-norm of the Haar-transformed time series. This again encourages stepwise time courses, which can be represented by a small number of Haar wavelets.

**Figure 1.**
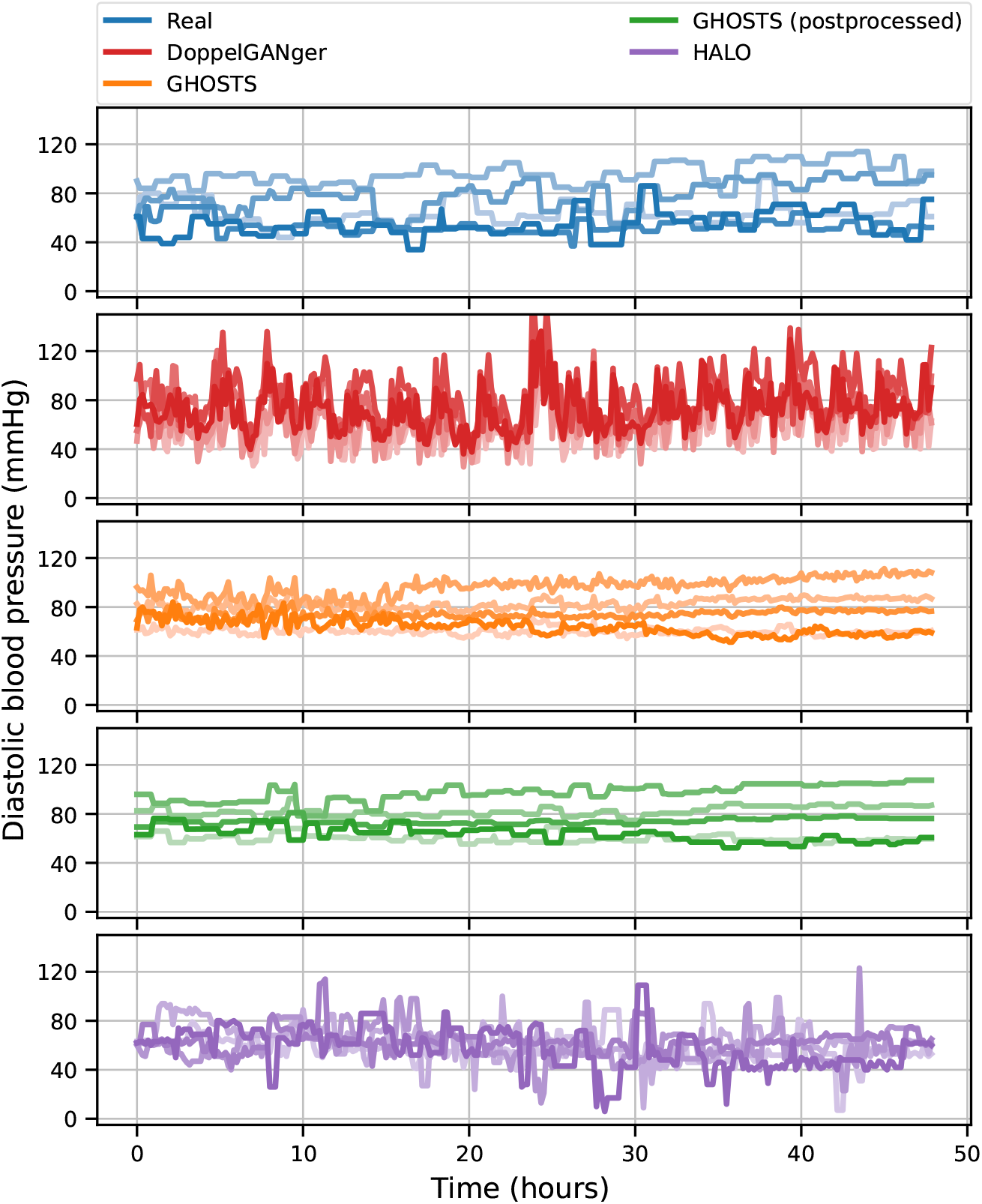
Characteristics of real intensive care unit (ICU) time series data compared to synthetic data generated by the DoppelGANger method as well as GHOSTS with and without postprocessing (GHOSTS_RAW and GHOSTS_POST). Real data were obtained from the MIMIC-IV database and used to train the GHOSTS and DoppelGANger models. Examples of real and synthetic diastolic blood pressure time series. Curves with different color tones correspond to independent samples.

The GHOSTS architecture consists of the generator *G*_*θ*_ and the discriminator *D*. The generator consists of two neural networks. The first one is a static attribute generator implemented as a multi-layer perceptron (MLP) with three dense layers, ReLU activations, and a batch normalization layer. The second one is an LSTM network acting as a time series generator. In analogy to the DG architecture, the discriminator consists of two independent networks: a general discriminator and an attribute-only discriminator. The general discriminator is applied to the combination of the attribute and time series, and the attribute discriminator is applied only to the static attributes. Both are MLPs with five dense layers, ReLU activations, and a minibatch discrimination bypass layer.

Since we observed mode collapse during the development of GHOSTS, we equip the discriminator with minibatch discrimination (MBD) layers as proposed by Salimans et al. (2016). The idea of MBD is to provide information about the samples in a batch to the discriminator and to penalize a lack of diversity in the batch. Minibatch discrimination is practically implemented as a layer parallel to the discriminator accepting the same input, but the outputs of the minibatch discrimination layer are concatenated with the output of the discriminator and passed to the final linear layer. We train GHOSTS using the Adam optimizer and reuse hyperparameter settings proposed for DoppelGANger (Lin et al., 2020a). Prior to training, all features are scaled to values between 0 and 1 using a maximum absolute scaling scheme. This scaling is reversed before the generated data are returned. All hyperparameters are set via hyperparameter optimization (HO). Methodological details as well as final parameters values are presented in Section S2.

##### Postprocessing

We observed that, despite the structural extensions to the existing DG methods described above, the generated data still contained subtle structural deviations from the real data. Real data contain certain regularities that are difficult to enforce with loss functions alone. Specifically, different physiological measures are constrained to attain only a finite number of different values due to quantization effects induced by the measurement process. Moreover, time series of distinct physiological quantities are also characterized by a finite number and characteristic distribution of sampling intervals. Accounting for these observations we devised the following routine to align the distributions of raw feature values and sampling intervals produced by GHOSTS with the empirical training distributions.

1. Each feature value in the generated time series is replaced by the closest value from the featurewise set of discrete values.
2. For each feature, the empirical distribution of the sampling intervals is estimated from the training data using a histogram with equal bin width of 10 minutes.
3. For each feature of each generated sample, new sampling points are defined by drawing randomly from the empirical distribution estimated from the training data in step 2.
4. The generated time series are downsampled to the sampling points defined in step 3 by taking the values at the sampling point and forward filling until the next sampling point.

#### 2.5. Classifier architecture

A deep neural network classifier 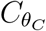 is used to distinguish real from generated data in the context of faithfulness assessment as well as to distinguish low from high SOFA scores as a downstream task in the context of utility assessment. The classifier network takes time series data **d**_time_, static attributes **d**_attr_ and descriptive summary statistics derived from the time-series data, *q*_*i,f*_ (**d**_time_) ∈ {0.2, 0.5, 0.9}, corresponding to the same ICU stay as input and outputs a continuous value 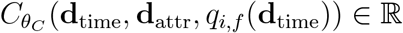 that can be interpreted as the probability that the ICU patient has a high SOFA score. Prior to training, all features are scaled to values between 0 and 1 using a standard scaling scheme.

The classifier architecture, refered to as MLP-LSTM-Static, combines an LSTM network, which processes the time series data, with an MLP network acting on static attributes and summary statistics derived from the same time series. The overall architecture consists of three parts: an LSTM layer with dropout, a sequence of dense (MLP) layers, and a last dense layer. First, the time series input is passed to the LSTM layers. In parallel, corresponding dense layers accept the concatenated static attributes and summary statistics. The output of the last time step of the last layer of the LSTM is concatenated with the output of the dense layers and passed to the last dense layer after which the logistic loss is applied. The numbers of LSTM and dense layers, the number of nodes of the LSTM and dense layers, and the dropout rate are hyperparameters that are optimized. The experimental details about utility assessment are described in Section S3.

#### 2.6. Performance metrics

The generation of synthetic hospital time series is a relatively novel field. When assessing the quality of generated data, their nature as time series with characteristic spatio-temporal dynamics needs to be taken into account, which requires dedicated performance metrics. Here, we discuss quantitative metrics to assess the faithfulness of generated data with respect to the training distribution as well as their utility with respect to a binary downstream classification task.

##### Faithfulness and diversity

Comparing the distributions of such high-dimensional hospital time series is non-trivial, especially in the presence of rich spatio-temporal correlations within and between features. Here, we evaluate the closeness of distributions up to second-order moments, capturing the faithful reproduction of spatiotemporal correlations in the generated data.

###### Similarity of raw feature values

We evaluate the similarity of the univariate distributions of the values of individual features in the real and synthetic training data by calculating the Wasserstein distance (WSD, Ramdas et al., 2017) between these distributions. In this univariate case, the WSD between probability density functions reduces to the total area between the corresponding cumulative distribution functions (CDF, c.f., Ramdas et al., 2017; Lipp and Vermeesch, 2023). Since different time series features have distinct ranges, we normalize Wasserstein distances per feature by the difference between the attained maximum and minimum values. Normalized WSD (WSD_norm_) is calculated based on empirically estimated CDFs (eCDFs).

Let 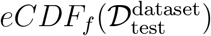 and 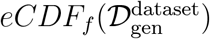 be eCDFs of one of the features *f* ∈ *F* in real and generated data respectively, where *eCDF*_*f*_ is an empirical cumulative distribution function 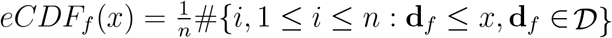 (here for convenience we use *eCDF*_*f*_ (𝒟) as shorhand for *eCDF*_*f*_ (*x*) defined for data 𝒟) and dataset MIMIC, eICU, then we define the normalized Wasserstein distance, ranging between 0 and 1, between them as

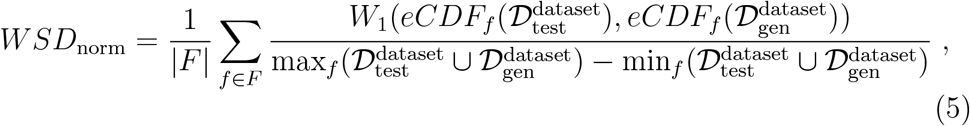

where

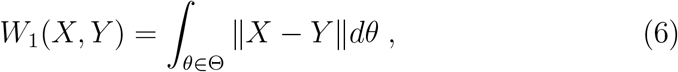

for arbitray cumulative distribution functions *X* and *Y*, and where max_*f*_ and min_*f*_ are the maxima over the samples *n* ∈ *N* and time indices *t*_*i*_ ∈ *T* over both datasets for feature *f* ∈ *F*.

###### Similarity of second-order statistics

We also measure the similarity of second-order statistics of real and generated data. Based on feature-by-feature correlation matrices, we define the following metrics: adapted correlation accuracy (CorrAcc, Tao et al., 2021; Li et al., 2023), mean absolute difference (CorrMAD, Li et al., 2023), mean squared error (CorrMSE), and mean Wasserstein distance (CorrWSD). In addition, we measure the reproduction of auto- and cross-spectral statistics based on lagged auto- and cross-correlation matrices, giving rise to two further families of metrics measuring deviations in autocorrelation (ADMAD, ADMSE, ADWSD), and deviations cross-correlation (CCDMAD, CCDMSE, CCDWSD), respectively. Here we mainly focus on divergence metrics based on the Wasserstein distance (CorrWSD, ADWSD, CCDWSD), while metrics comparing distribution means (CorrMAD, CorrMSE, ADMAD, ADMSE, CCDMAD, CCDMSE) are presented in Supplementary Section S4. The pairwise Pearson correlation coefficient (PCC) of the *n*^th^ sample is defined as

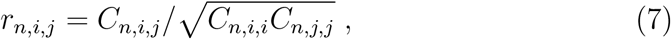

where

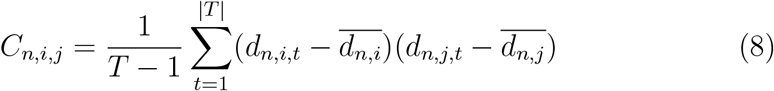

and

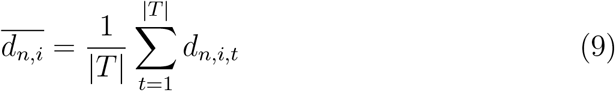

are empirical estimators of feature covariances and means, respectively. Here, *d*_*n,i,t*_ is an entry in matrix **d**_time_ = (*d*_*n,f,t*_) corresponding to the *n*^th^ sample, where *f* ∈ *F* and *t* ∈ *T*. Let *K* be the set of all pairs (*i,j*) of features in *F*, 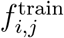 is the discretized correlation between features *i* and *j* in the real data, and 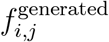 is the corresponding value of the generated data, then correlation accuracy (CorrAcc, Tao et al., 2021) is the fraction of matching correlations over |*K*| pairs, see Equation (10). We discretize the correlation as follows: mean PCC for each feature pair is defined as 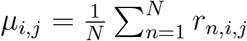 and discretized into seven bins: low, medium, and high positive correlation (*µ*_*i,j*_ ∈ [.1, .3), *µ*_*i,j*_ ∈ [.3, .5), *µ*_*i,j*_ ∈ [.5, 1], respectively), low, medium, and high negative correlation (defined analogously), and no correlation (*µ*_*i,j*_ ∈ (−.1, .1)):

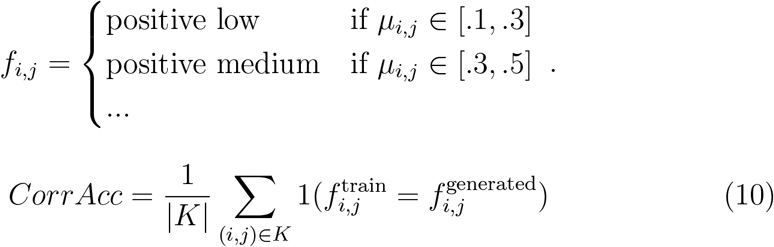

Let 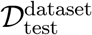 and 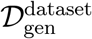 be datasets in real and generated data respectively, where dataset ∈ {MIMIC, eICU}, then 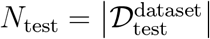 and 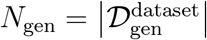. We also define 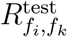 and 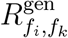 to be the empirical cumulative distribution functions of the the correlation coefficients 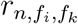 computed on test and generated sets, respectively.

To obtain a metric that is more sensitive to minor deviations in PCC than CorrAcc, we also compute the mean Wasserstein distance between PCCs observed in training and generated data:

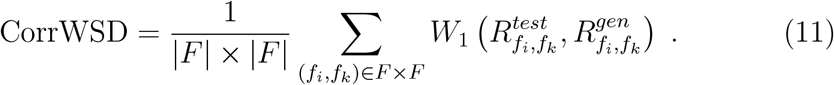

We further define the autocorrelation of the time series *d*_*n,f,t*_ of feature *f* in sample *n* and at lag *h* as

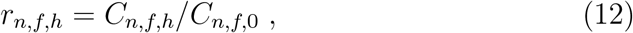

where *C*_*n,f,h*_ is the estimated autocovariance and *R*_*f,h*_ denotes the eCDF of the autocorrelation coefficients *r*_*n,f,h*_.

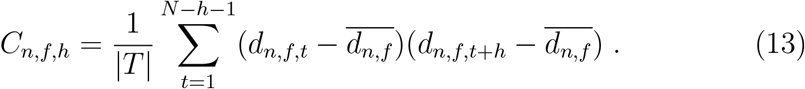

Analogous to quantities defined for PCC, we define the ACWSD metric for assessing the reconstruction of autocorrelation properties:

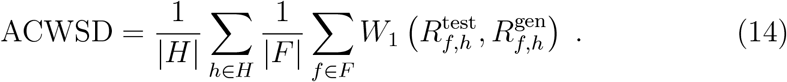

Finally, we define the cross-correlation of the time series for selected lags *h* ∈ *H*, |*H*| ≤ |*T* | as

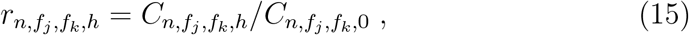

where

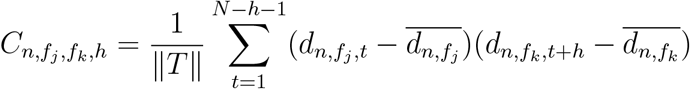

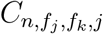 is the estimated cross-covariance. Further, we define 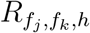 to be the eCDF of the cross-correlation coefficients 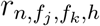. Then, the CCWSD metric for assessing cross-correlation reconstruction is defined as:

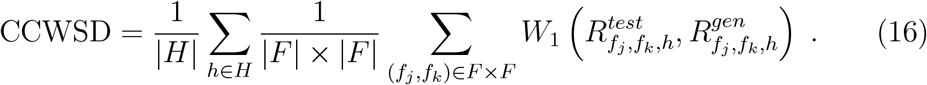

###### Similarity of attributes

We define an analogous faithfulness metric for static attributes as

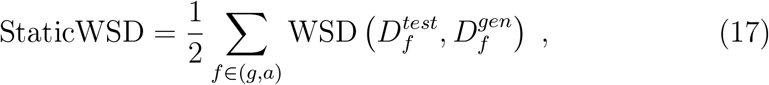

where *d*_*n,f*_ is an element of **d**_attr_ = (*d*_*i,f*_) where *n* ≤ *N* and *f* ∈ (*E*(*g*), *a*), where *g* ∈ {*F, M*} is biological sex, and *a* ∈ ℕ^+^ is the patient age at the time of ICU admission and *D*_*f*_ is the eCDF of the observed *d*_*i,f*_.

###### Discriminability

We also train classifiers to distinguish between real and synthetic samples separately for each generative model. We measure the discriminability of these classifiers as a further metric of distributional similarity. Details about architecture can be found in Section 2.5 and details about training in Section S3

The performance of the classifier is evaluated using the area under the receiver operating characteristic curve (AUROC) and the area under the precision-recall curve (AUPRC). The ROC curve depicts the true positive rate (TPR) as a function of the false positive rate (FPR) over all possible classifier thresholds (Fawcett, 2006). Analogously, the precision-recall curve illustrates the trade-off between precision and TPR (Davis and Goadrich, 2006). The AUROC is computed by numerically approximating the integral under the ROC curve, aggregating the trade-off between TPR and FPR in a single number, while, analogously, AUPRC is computed by approximating the integral under the precision-recall curve:

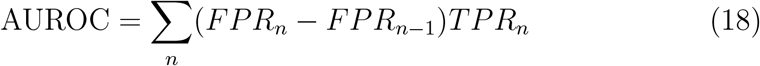

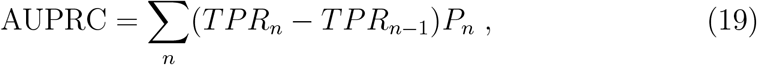

where *FPR*_*n*_, *TPR*_*n*_, and *P*_*n*_ are FPR, TPR, and precision at the n^th^ threshold, and where the number of thresholds is equal or less than the number of samples. For binary classification tasks, the performance of a random classifier is characterized by AUROC = 0.5 and 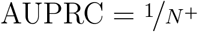, where *N* ^+^ is the number of positive test samples.

##### Privacy

To evaluate the privacy-preserving capability of the generative models, we perform a membership inference attack developed by Chen et al. (2020). It is formulated as estimating *P* (*m*_*i*_ = 1|*x*_*n*_, *θ*), the probability of a sample *n* being a member of the training dataset, given model weights *θ*. We assume that this probability is proportional to the probability that the sample is generated by the model, *P* (*m*_*n*_ = 1|*x*_*n*_, *θ*) ∝ *P*_*G*_(*x*|*θ*). We perform the version of the attack in a complete black-box setting, where the attacker only has access to the generated samples but not to the model itself. In this attack, *P*_*G*_(*x*|*θ*) is approximated by

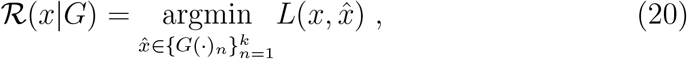

where *L* is a distance metric and 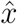 is a sample generated by *G*(·). In our experiments, *k* is equal to the training size (*N*_train_). The ability of ℛ (*x*|*G*) to discriminate between training data and data not used in training is then evaluated using the AUROC metric.

##### Utility

To assess the utility of the synthetic data in a realistic clinical prediction setting, we consider a binary classification problem, where the prediction target is whether the SOFA score above or below a certain threshold. 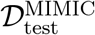 is further split into training and testing sets for the downstream task. Analogous datasets are generated using generative models. Then, classifiers are trained using real and synthetic training and test datasets to output the corresponding SOFA score labels (outputs). Only samples with either high (SOFA^+^) or low (SOFA^-^) SOFA score are included, defining a binary classification task. More details can be found in Section S3.

Two scenarios are considered: data utility and data augmentation. Data utility refers to the case where a model is trained on a dataset from one domain and its performance is assessed on a dataset from another domain.

We investigate all four combinations of training a model on either real or synthetic training data and testing the model on either real or synthetic data test data. These combinations are referred to as ‘train on synthetic, test on real’ (TSTR), ‘train on real, test on real’ (TRTR), ‘train on synthetic, test on synthetic’ (TSTS), and ‘train on real, test on synthetic’ (TRTS). Data utility is defined as the performance of a model trained on synthetic data when applied to real data (TSTR case), whereas the TSTS and TRTR cases provide upper bounds of the performance to be expected.

In the data augmentation case, the model is trained on a combination of real and synthetic data and applied to real test data, where the amount of synthetic data to be used is a parameter.

Test performance is measured using the area under the receiver operating characteristics curve (AUROC) and the area under the precision-recall curve (AUPRC), see Section 2.6.

## 3 Results

All experiments reported here were performed on a workstation with 128 cores, 256 GB of memory, and an A100 (40GB memory) GPU. Computation of the complete experiments took approximately 7 days.

We trained the GHOSTS, DG and HALO models on the 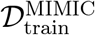 dataset as outlined for GHOSTS in Section 2.4. For DG and HALO, we use the authors’ implementations (Lin et al., 2020b; Theodorou et al., 2023) with their default parameters. Next, we sampled *N*_train_ = 11, 996 and *N*_test_ = 4, 220 samples using each model, preserving the class proportions of 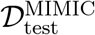 with rejection sampling. The quality of the generated data was assessed in terms of their faithfulness, diversity, privacy, and utility with respect to SOFA score classification using the metrics outlined in Section 2.6.

Table 1 summarizes the performance of all methods with respect to how faithful properties of the real data such as raw values, auto- and cross-correlations are reflected in the generated data. In addition, the degree of privacy preservation under MIA is presented. The table focuses on faithfulness metrics employing the WSD to measure divergences between the distributions distributions of these quantities, whereas metrics quantifying differences in central tendencies are presented in Table S8. Standard errors are estimated using the bootstrap method (K=1,000). Across almost all metrics, GHOSTS achieves either superiority over DG and HALO, or comparable results. Overall, GHOSTS_RAW outperforms DG in all metrics and is on par in terms of the privacy metric, with GHOSTS_POST outperforming DG and HALO in the same metrics. These results indicate that both GHOSTS variants represent a marked improvement compared to DoppelGANger and HALO, although the postprocessing does not seem to lead to a more faithful representation of spatio-temporal dynamics. On the other hand, the HALO model outperforms the GHOSTS_RAW model in terms of autocorrelation and cross-correlation faithfulness.

**Table 1:** Quantitative evaluation of how faithfully synthetic MIMIC-IV data generated by DoppelGANger, GHOSTS without postprocessing (GHOSTS_RAW) and GHOSTS with postprocessing (GHOSTS_POST) reproduce the distribution of real MIMIC-IV data. WSD_norm_ refers to the normalized Wasserstein distance between univariate distributions of raw values of individual time series features presented in Equation (5). CorrWSD refer to the Wasserstein distance between distributions of real and synthetic |*F*| × |*F*| temporal Pearson correlations between features, corresponding to Equation (11). Similarly, AC_WSD, CC_WSD denote auto- and correlation similarities derived from |*F*| × |*F*| × *N*_lag_ lagged cross-covariance matrices, respectively, corresponding to Equations (14) and (16). Privacy refers to the Equation (20). Static_WSD denote comparisons between univariate distributions of static attributes presented in Equation (17). Discrimination AUROC and AUPRC refers to discriminaboloty as measured by area under receiver operator curve and area under precision-recall curve, described in Equations (18) and (19), respectively. Metrics are reported as mean ± standard error of the mean, and ↓/↑ indicate that lower/higher values are better, respectively. Uncertainty was estimated using bootstrap method (K=1,000)

|  |  | GHOSTS_RAW | GHOSTS_POST | DoppelGANger | HALO |
| --- | --- | --- | --- | --- | --- |
| | Static WSD ↓ | **0.4120 ± 0.3935** | **0.4120 ± 0.3935** | 1.4100 ± 1.4019 | 2.6360 ± 2.5141 |
| | $WSD_{\text{norm}}$ ↓ | **0.0038 ± 0.0022** | 0.0040 ± 0.0031 | 0.0142 ± 0.0098 | 0.0105 ± 0.0032 |
| | CorrWSD ↓ | 0.1120 ± 0.0003 | **0.1029 ± 0.0003** | 0.1854 ± 0.0004 | 0.1130 ± 0.0004 |
| | AC WSD ↓ | 0.0801 ± 0.0422 | 0.0538 ± 0.0413 | 0.1383 ± 0.0485 | **0.0463 ± 0.0188** |
| | CC WSD ↓ | 0.1304 ± 0.0002 | **0.1107 ± 0.0003** | 0.2490 ± 0.0003 | 0.1219 ± 0.0004 |
| Discrimination ↓ | AUROC | 0.9898 ± 0.0009 | **0.6580 ± 0.0058** | 1.0000 ± 0.0000 | 0.9580 ± 0.0021 |
| | AUPRC | 0.9891 ± 0.0014 | **0.6416 ± 0.0068** | 1.0000 ± 0.0000 | 0.9487 ± 0.0039 |
|  | Privacy | **0.5019 ± 0.0004** | **0.5035 ± 0.0005** | **0.5041 ± 0.0004** | **0.5019 ± 0.0006** |

Figure 2 compares real MIMIC-IV and synthetic GHOSTS data distributions across |*F*| = 9 clinical variables. GHOSTS overall successfully reproduces the MIMIC-IV distributions. The sharp peaks in the MIMIC dataset correspond to imputed median values and are, notably, not as accurately reproduced by GHOSTS as the rest of the distributions. Similar results are observed for postprocessed GHOSTS, see Figure S2. On the other hand, DoppelGANger and HALO fail to capture the mode of systolic blood pressure, heart rate, and oxygen saturation, respectively, see Figure S3 and Figure S4. Notably, HALO reproduces imputation peaks only in some variables, like glucose (overestimates the peak at least two times) and diastolic blood pressure, but not in oxygen saturation. GHOSTS and DG seem to avoid the reproduction of imputation peaks. Moreover, other variables, such as SOFA and systolic blood pressure are unfaithfully generated by HALO. For the SOFA score, HALO created a non-existent bimodal distribution. The generated systolic blood pressure distribution is skewed to the left. Making similar comparison on the smaller eICU dataset, we observe that GHOSTS represents all variables consistently well (see Figures S5 and S6). On the other hand, DoppelGANger and HALO fail to capture the distribution of diastolic blood pressure, see Figures S7 and S8.

**Figure 2.**
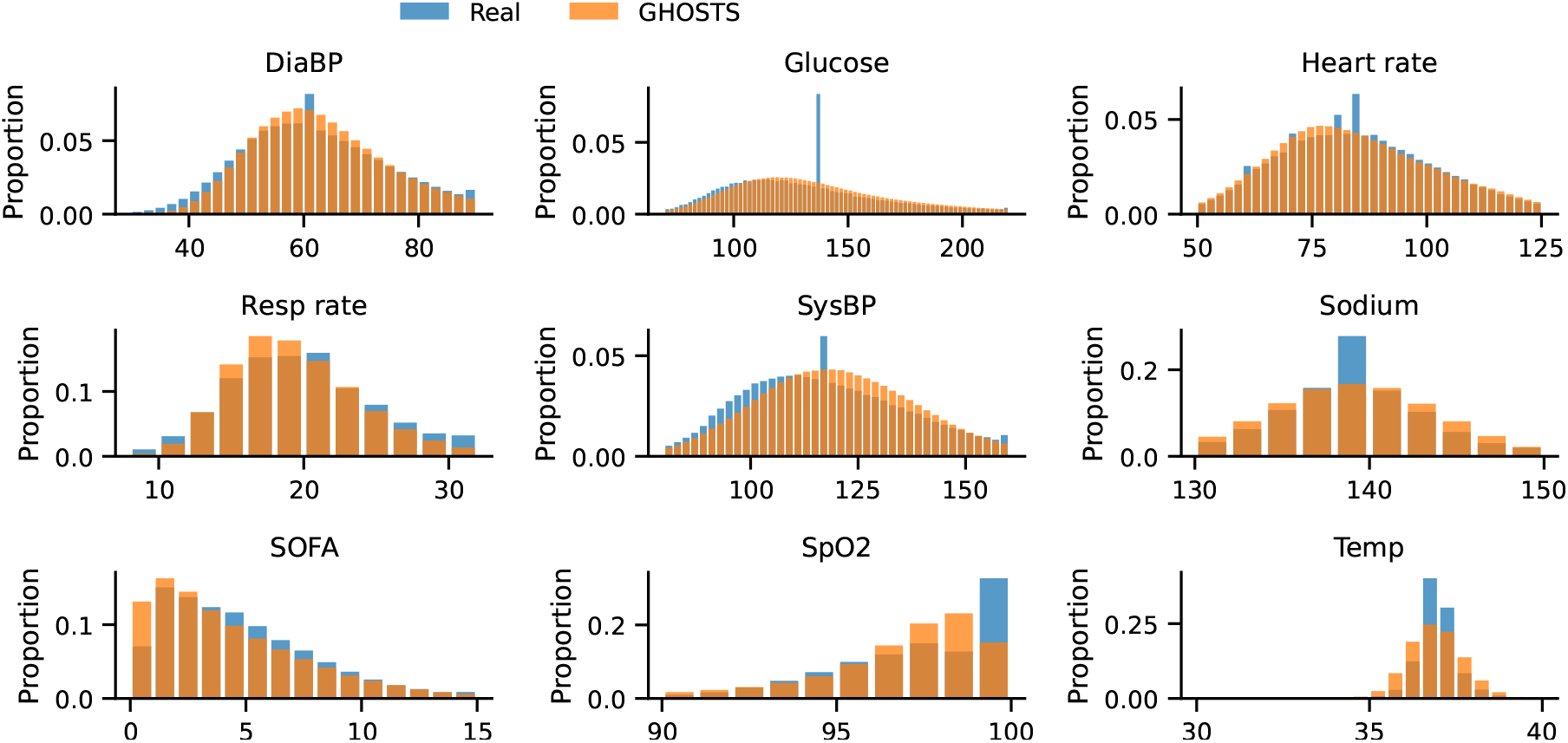
Comparison of the distributions of |*F*| = 9 clinical variables between real MIMIC-IV data and synthetic GHOSTS data. The aggregated statistics are normalized to sum to 1. Abbreviations: DiaBP – diastolic blood pressure, Resp rate – respiratory rate, SOFA – Sequential Organ Failure Assessment (SOFA) score, SpO2 – oxygen saturation, SysBP – systolic blood pressure, Temp – temperature. Apart from peaks related to data imputation, which are not reproduced by GHOSTS, a strong resemblance between real and synthetic data distributions is observed for all features.

Figure 3 A depicts the utility of the synthetic data for the SOFA score classification task, which is operationalized as the performance of the MLP-LSTM-Static classifier and measured using the AUPRC and AUROC metrics. Blue color marks the performance of the baseline model, which was trained and also tested on real MIMIC data. The x-axis shows the performance of models trained on real data and tested on synthetic data generated by different models. Conversely, the y-axis shows the performance of the model trained on synthetic and tested on real data. Each colored circle represents the performance of these classifiers on one bootstrap sample drawn from the respective test sets. Dashed curves and colored crosses correspond to classifiers trained and tested on synthetic data (TSTS). Training and testing on synthetic GHOSTS data leads to high performance, which falls only slightly behind models trained and tested on real data. This indicates that GHOSTS learned to generate discriminative features. In contrast, in the TSTS setting, DoppelGANger outperforms models trained and tested on the real data, suggesting it augments the feature space with highly discriminative features not found in the real data. The TSTS performance of the HALO model is inferior to that of GHOSTS. The performance is similar for models trained on real MIMIC data (TRTS settings) and improves moderately in the relevant setting in which models are tested on real MIMIC data (TSTR setting), except for HALO, which is outperformed by GHOSTS and DoppelGANger in both settings. Notably, GHOSTS with and without postprocessing substantially outperforms DoppelGANger in all settings and with respect to the AUROC metric. The reader is refered to Table S5 for a tabular summary of the same results.

**Figure 3.**
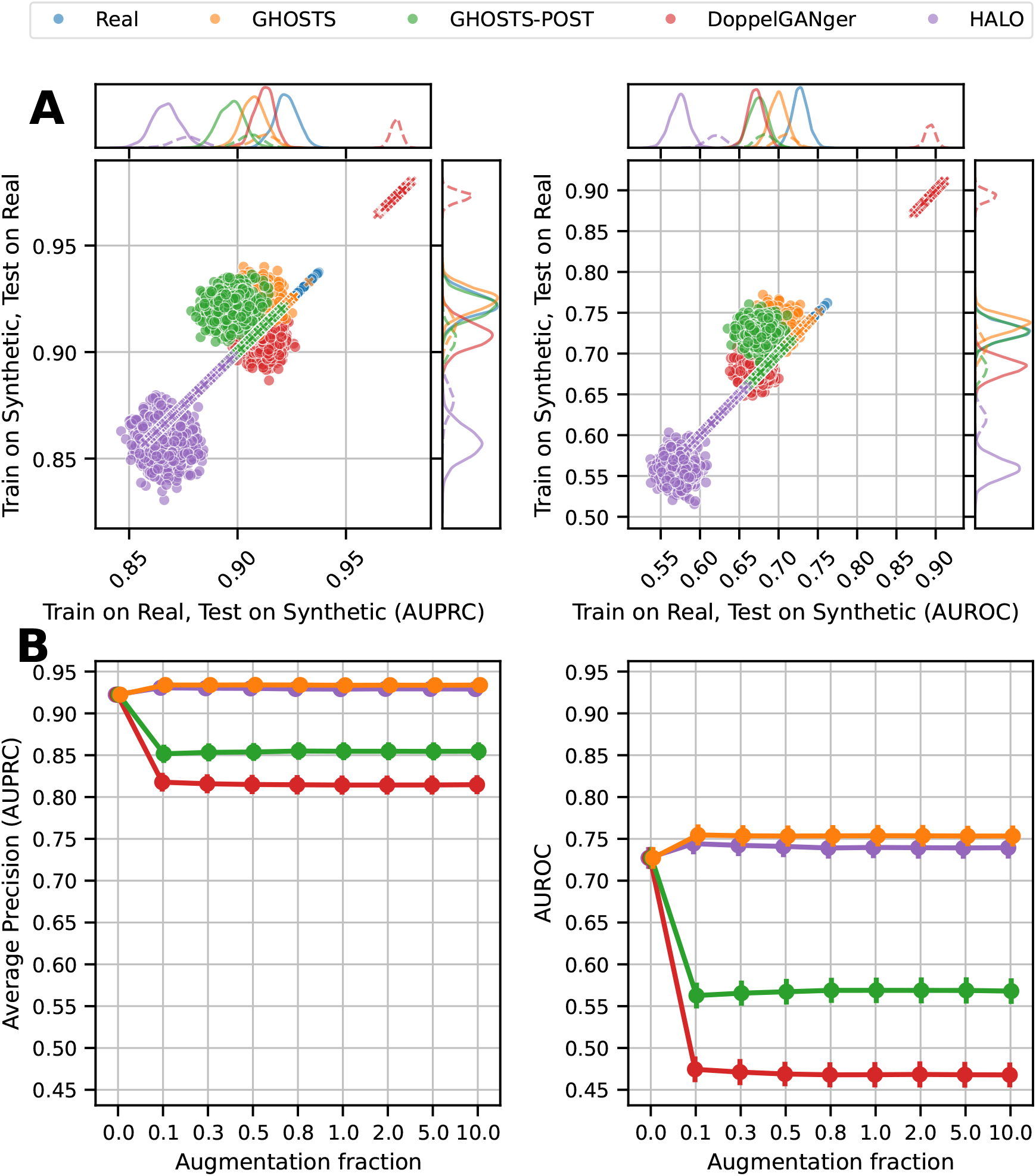
*(previous page)*: Performance of MLP-LSTM-Static classifiers in the downstream SOFA score classification task across synthetic data generated by four methods, and the real data as a baseline. Two performance scores are evaluated: Left – area under the precision-recall curve (AUPRC), calculated as average precision; right – area under receiver operator curve (AUROC). **Panel A** shows the classifier performance in the data utility scenario for four distinct settings: *Train on Synthetic, Test on Real* (TSTR), *Train on Real, Test on Synthetic* (TRTS), *Train on Synthetic, Test on Synthetic* (TSTS), and *Train on Real, Test on Real* (TRTR). In each setting, a classifier was trained either on real data or synthetic data generated by one of the considered methods, and evaluated on separate test data, wich are again either real data or synthetic data generated by the same model. Scatterplots show bootstrapped test performance, where each dot is the performance of the classifier on a bootstrapped version of the test set. Density curves on the sides show marginal distributions. Yellow, green, red, and purple circle markers show the performance of GHOSTS, GHOSTS-POST, DoppenGANger, and Halo in the TSTR and TRTS scenarios. Dashed lines and cross markers correspond to the TSTS (yellow, green, and red colors) and TRTR (blue color) scenarios. GHOSTS and GHOSTS-POST achieve highest performance in the TSTR setting most relevant for utility evaluation. **Panel B** illustrates classification performance in the data augmentation setting. Classifiers were trained on a mixture of real and synthetic data, and then tested on the hold-out set of baseline data. Augmentation fractions correspond to the fraction of the synthetic data that are added to the baseline data. E.g. the size of the training set for the augmentation fraction 0.5 is 1 + 0.5 = 1.5 × |*D*_baseline_|. Hence, the augmentation fraction 0 corresponds to the training of the classifier without data augmentation. GHOSTS and HALO achieve highest performance across all augmentation fractions. However, no systematic increase in performance is observed as a result of adding more synthetic data.

Figure 3 B illustrates the utility of synthetic data in the data augmentation task, which is expressed as the performance of the MLP-LSTM-Static classifier trained on mixtures of real and synthetic data, and then tested on real hold-out data. Augmentation fractions correspond to the fraction of generated data added to the real training data. GHOSTS demonstrates superior performance across all metrics and augmentation fractions over DoppelGANger and slight improvements over HALO, see Table S6 for detailed results.

Table S9 summarizes the results for different faithfulness metrics on the eICU dataset in analogy to results reported for MIMIC-IV. Across all except cross-correlation metrics, GHOSTS achieves superiority over DoppelGANger. Overall, comparing these results with Table 1 and noting that the training split of the eICU dataset is around 4 times smaller than that of the MIMIC-IV dataset, we observe that GHOSTS scales better in correlation and cross-correlation metrics, while DG demonstrates very moderate or no improvement when trained on larger MIMIC-IV dataset.

Figure 4 shows the change of faithfulness as the training set size increases for GHOSTS. According to almost all time series metrics, there is a significant improvement as the training set size increases, with normalized Wasserstein distances between univariate distributions of raw values showing the most significant improvements. See Table S7 for a tabular summary of the same results.

**Figure 4.**
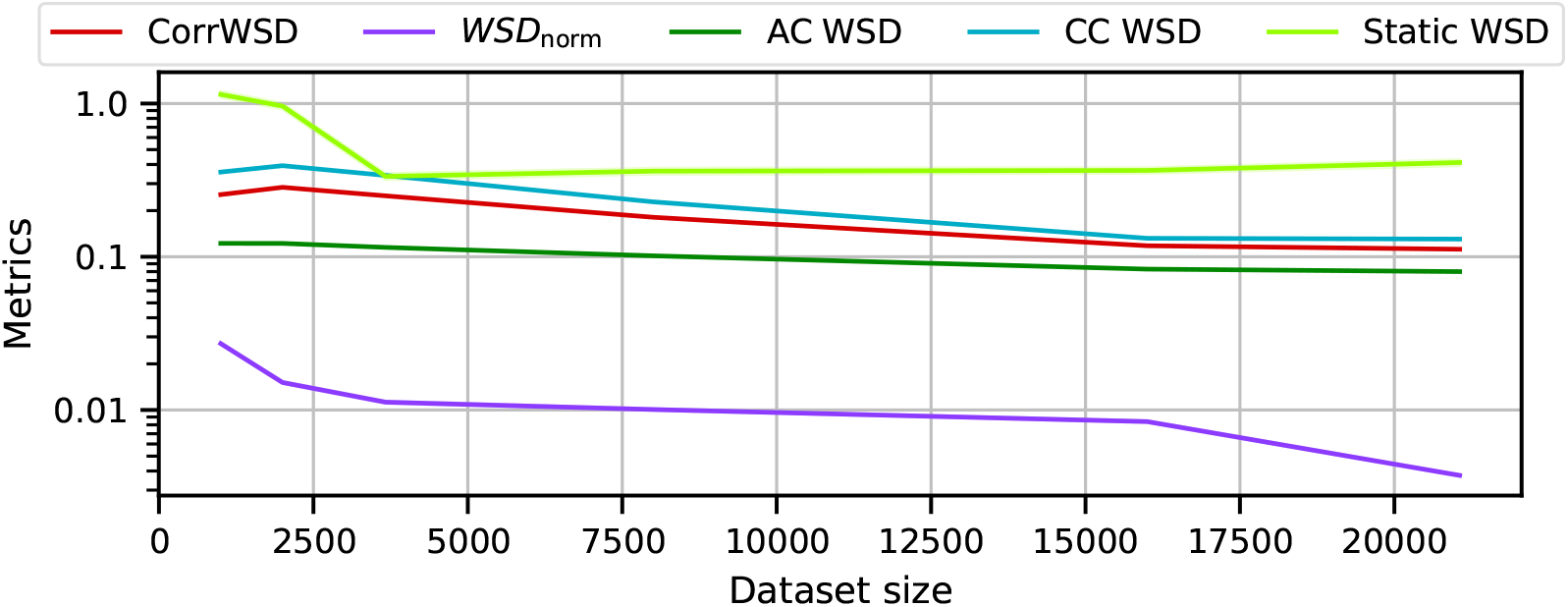
Comparison of metrics across different training set sizes 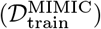 for GHOSTS. Abbreviations: CorrWSD - Wasserstein distance between distributions of real and synthetic temporal Pearson correlations corresponding to Equation (11), *WSD*_norm_ - normalized Wasserstein distance between univariate distributions of raw values presented in Equation (5), AC WSD and CC WSD refer to auto- and cross-correlation similarities referring to Equations (14) and (16), Static WSD - comparison between univariate distributions of static attributes from Equation (17)

## 4. Discussion

We presented GHOSTS, a novel generative model architecture tailored to generate realistic EHR data comprising heterogeneous longitudinal and tabular data types. Previous methods are either unable to generate medical time series data, such as data collected in ICUs, or tend to generate unrealistic data with even sampling patterns and generated values that are frequently outside of the range of real training data. Moreover, EHR data generated by existing methods often shows signs of mode collapse, indicating convergence issues during model training. Through the introduction of novel loss functions and postprocessing routines, GHOSTS overcomes these limitations and is able to generate unevenly sampled realistic ICU time series. We trained and validated GHOSTS on a large patient cohort extracted from the MIMIC IV and eICU databases. The results indicate a significant gain over the DoppelGANger and HALO models, both in terms of faithfully reconstructing the distributions of essential features of the MIMIC data and in terms of the classification performance in a clinical downstream classification task.

We note that data modelling technique, employed in our model, of stepwise constant functions is functionally equivalent to a value-timestamp format, where each data point is characterized by its corresponding value and the time at which it was recorded. This representation preserves the temporal dynamics of the data and enables the model to effectively capture sequential dependencies.

We observed that DoppelGANger is not well suited for EHR data generation as it is restricted to evenly sampled data. While unevenly sampled data can be resampled to be applicable to architectures like DG, this leads to stepwise behavior, which is difficult for the majority of generative models, including DG, to reproduce (c.f. Figure 1). Moreover, we observed that DG generates values outside of the range of the reference data. To remedy these issues, we designed novel loss functions as well as a postprocessing procedure. Our quantitative and qualitative analyses indicate that both solutions successfully led to the generation of more realistic, faithful, and useful data.

While HALO shows adequate performance, the data modelling technique that it is based on limits its ability to faithfully generate distributions of values. For instance, Figure S4 shows that certain clinical variables, such as glucose, that contain significant amount of missing samples, therefore have large peaks. We notice that HALO exagerates the imputation peak while generating synthetic ICU stays. Moreover, this exaggerated peaks can be seen in other clinical time series, like diastolic blood pressure, systolic blood pressure, heart rate, and sodium. Provided that electronic health records often contain missing samples, it makes it difficult to recommend HALO as a suitable data generation method for health data. Another obesrvation of HALO is that it tends to generate short ICU stays despite the training data having the ICU stays of the same length. For our experiments we needed to generate significantly more (x10) synthetic ICU stays and then reject the shorter ones.

We also observed that the distribution of generated values becomes more similar to that of the reference dataset when the proposed postprocessing routine is applied. However, it is noteworthy that this also adversely affects auto- and cross-correlations to a certain degree. Notwithstanding, performance in the clinically relevant classification task demonstrates that the magnitude of this influence is negligible. One possible explanation is that the smoothing applied during postprocessing is too high. By smoothing noisy data points, post-processing improves the eCDF score but degrades other faithfulness metrics. However, since classification is less dependent on individual noisy measurements but rather takes into account the entire temporal trajectory of the data, the impact of postprocessing on classification performance is negligible.

All methods show low performance in the data augmentation task. One possible explanation is a ceiling effect where the classifier already performed optimally in the baseline TRTR setting in terms of an optimal relation between model complexity and training dataset size. In this scenario, adding generated data would not lead to a helpful regularization effect.

As part of the numerical benchmarks conducted in this work, we trained GHOSTS on the large MIMIC IV and eICU datasets, thereby creating synthetic MIMIC IV and eICU patient cohorts. We will publish these synthetic datasets, the trained MIMIC-GHOSTS and eICU-GHOSTS models as well as all code associated with data extraction, pre- and postprocessing, model design and training, and performance evaluation upon manuscript acceptance. We expect that the public availability of these tools will promote machine learning research in the ICU field and lead to the development of novel methods and models. Specifically, we expect GHOSTS to enable in-silico clinical trials, where it is necessary to generate synthetic data for separate control and treatment groups. GHOSTS already has a built-in mechanism to condition its output on arbitrary static attributes. Thereby, it is able to generate counterfactual patient trajectories that can be used to estimate treatment effects (Myles et al., 2023). This is especially relevant in intensive care, where clinical trials frequently cannot be conducted due to ethical concerns.

GHOSTS is still under refinement, though. One current observation is that, while the devised postprocessing routine does seem to improve the overall faithfulness of the raw data value distribution, it also seems to adversely affect the reconstruction of spatio-temporal dynamics encoded in the auto- and cross-spectra of the data. Notwithstanding, this result had negligible influence on the performance of ML models in a clinically relevant classification task.

Besides the method itself, this study also has limitations. While we tried to exhaustively measure the quality of the generated data with different performance metrics, we did not assess the realism of the generated data as perceived by human practitioners, which could be an important factor influencing the acceptance of ML models trained on synthetic data by clinicians. In future work, we plan to conduct studies with clinical experts to guide the assessment and further development of our model. We also plan to train GHOSTS on additional patient cohorts and to study the across-site transferability of clinical prediction models and the possible role of synthetic data to improve transferability. Finally, our quantitative evaluation focused on the aspects of faithfulness, diversity, and utility. While the design of GHOSTS effectively prevents mode collapse, there is no theoretical guarantee to exclude that the model may memorize and reproduce individual patients’ data. It is thus of utmost importance that synthetic data fulfill privacy requirements. This was assessed here for a single type of privacy attack. Future work will consider a wider range of attacks and investigate privacy-preserving frameworks such as differential privacy in the context of generative time-series modeling.

## 5. Conclusion

In summary, we have presented GHOSTS as the first generative model capable of generating realistic ICU times with associated static patient attributes. As demonstrated in a large ICU data corpus, data synthesized by GHOSTS accurately resemble real training data and preserve details such as artifacts introduced by uneven sampling intervals of different time series measurements. Moreover, data generated by GHOSTS can be used to train machine learning models to achieve competitive performance on real data in clinical prediction tasks. Future work will extend the GHOSTS-Bench suite to include a larger variety of privacy preservation performance metrics.

## Data Availability

All data produced in the present study will be available upon reasonable request to the authors

## Acknowledgements

This work has been performed within the “Metrology for Artificial Intelligence in Medicine (M4AIM)” programme funded by the German Federal Ministry for Economy and Climate Action (BMWK) in the frame of the QI-Digital Initiative. NG is funded by the German Academic Scholarship Foundation.

## S1. Feature ranges

**Table S1:** List of features extracted from the MIMIC-IV database. Min and Max columns refer to the minimal and maximal values of the raw extracted features. Valid min and max refer to the minimal and maximal values of the preprocessed data after outlier removal.

| Feature name | Min | Valid min | Valid max | Max | Unit |
| --- | --- | --- | --- | --- | --- |
| Diastolic blood pressure | 1.00 | 1.00 | 293.00 | 299.00 | bpm |
| Systolic blood pressure | 0.10 | 0.20 | 365.00 | 365.00 | bpm |
| Respiratory rate | 0.13 | 1.00 | 69.00 | 69.00 | bpm |
| Heart rate | 1.00 | 1.00 | 295.00 | 296.00 | bpm |
| SpO2 | 0.01 | 0.30 | 100.00 | 100.00 | % |
| SOFA score | 0.00 | 0.00 | 23.00 | 23.00 | point |
| Glucose | 0.12 | 33.00 | 1894.00 | 1 653 550.00 | mg/dL |
| Sodium | 1.36 | 77.00 | 186.00 | 1336.00 | mmol/L |
| Temperature | 15.00 | 26.10 | 42.30 | 43.11 | C |

## S2. Hyperparameter optimization

To select the hyperparameters of the GHOSTS and DoppelGANger models, we conducted hyperparameter optimization (HO) using the Optuna library (Akiba et al., 2019) in combination with tree-structured Parzen estimators (Bergstra et al., 2011), hyperband pruning (Li et al., 2017), and a reduction factor of 2. We used validation split *N*_val_ = 1, 282 described in Section 2.1 for HO. The optimization was run for around three days until the first 150 hyperparameters were tested, using the sliced Wasserstein distance (Bonneel et al., 2015) between the generated samples and the validation set as the performance metric. Then, we conducted an additional round of hyperparameter optimizations, where individual loss terms were separately iterated through a fixed linear grid of size 10. Parameters of the loss function *α, β*, and *γ* were iterated from 0.01 to 10, from 1.0 to 30.0, and from 1 to 40, respectively. The final set of hyperparameters was selected using the same criteria as in previous stage. The final regularization constants *α, β*, and *γ* were set to *α* = 0.22, *β* = 11.2, and *γ* = 25.7. The number of latent dimensions of the MBD layers was set to 7.

Due to the large number of hyperparameters in all models, we optimized only subsets of them, see Table S2 and Table S3, leaving various of them fixed for both architectures, see Table S4.

The HALO model was not hyperparameter optimized and default parameters were used, since the model was optimized on the MIMIC data in the original publication.

**Table S2:**
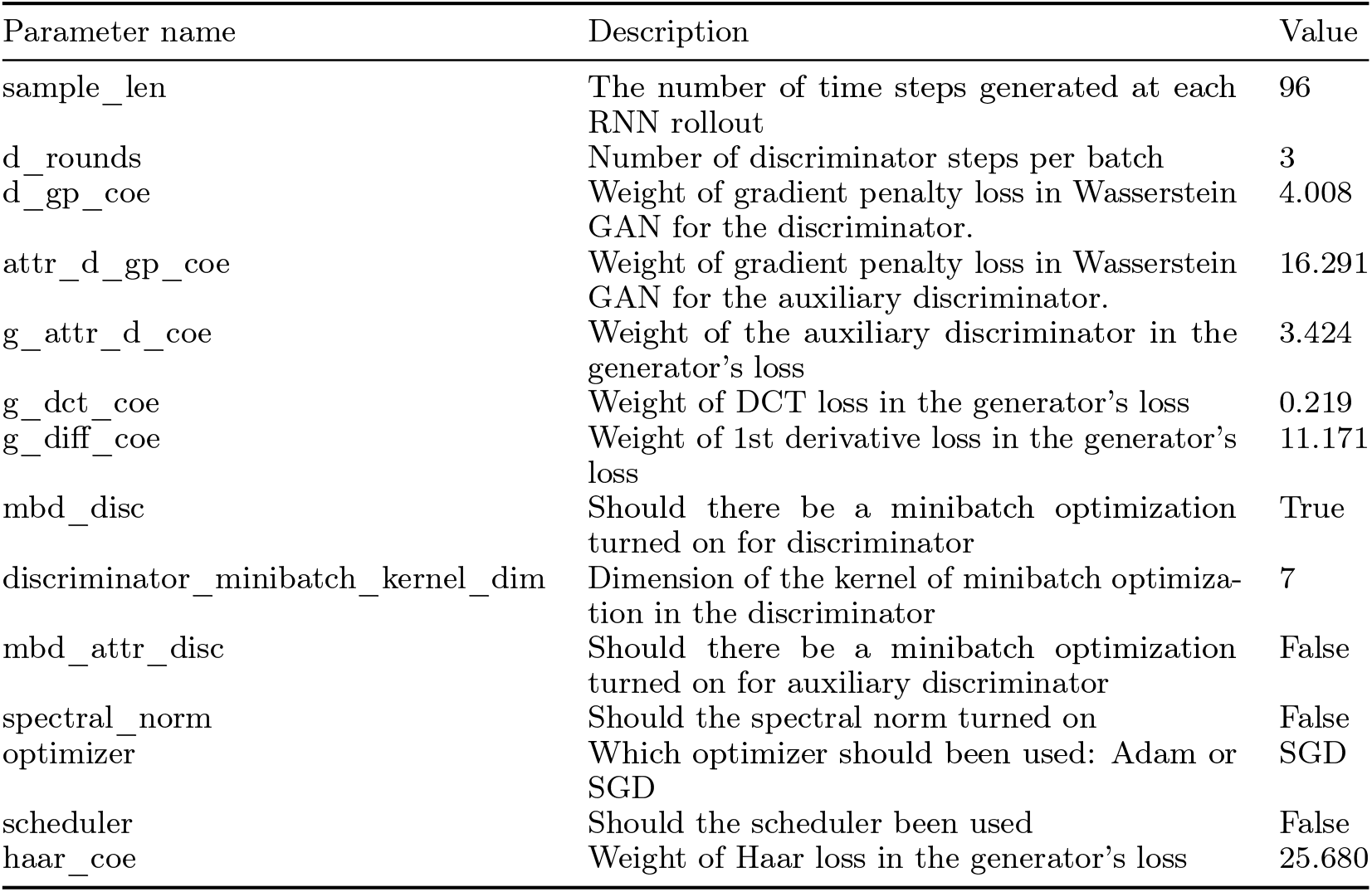
Hyperparameters optimized for GHOSTS.

## S3. Experimental details of the utility assessment

To test the performance of models trained on synthetic data in a down-stream classification task, the 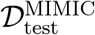 data were preprocessed to yield labels for the classification by partitioning *T* into the first 42 hours and the last 4 hours, discarding the 2-hour gap in between. The SOFA scores of the last 4 hours were used to create a label for the SOFA score class and the SOFA scores of the first 42 hours were removed from the data. Next, the test data were further divided into train, validation, and test splits, where we stratified patients by their SOFA score class (SOFA^-/+^) as defined below.

**Table S3:**
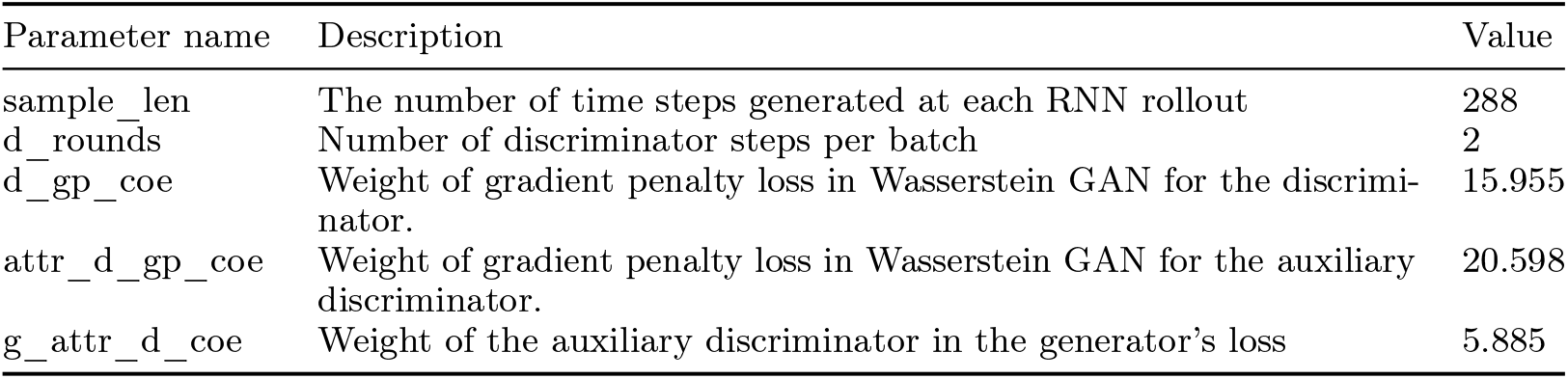
Hyperparameters optimized for DoppelGANger.

**Table S4:** Hyperparameters reported in DoppelGANger that were adopted for GHOSTS.

| Parameter name | Description | Value |
| --- | --- | --- |
| g_lr | The learning rate in Adam for training the generator | 0.001 |
| d_lr | The learning rate in Adam for training the discriminator | 0.001 |
| attr_d_lr | The learning rate in Adam for training the auxiliary discriminator | 0.001 |
| attribute_latent_dim | The dimension of noise for generating attributes | 5 |
| feature_latent_dim | The dimension of noise for generating features | 5 |
| generator_attribute_num_layers | The number of layers in attribute generator | 3 |
| generator_attribute_num_units | The number of latent units in attribute generator | 100 |
| generator_feature_num_layers | The number of layers in feature generator | 1 |
| generator_feature_num_units | The number of latent units in feature generator | 100 |
| discriminator_num_units | The number of latent units in discriminator | 200 |
| discriminator_num_layers | The number of layers in discriminator | 5 |
| aux_discriminator_num_units | The number of latent units in auxiliary discriminator | 200 |
| aux_discriminator_num_layers | The number of layers in auxiliary discriminator | 5 |
| d_beta1 | The beta1 in Adam for training the discriminator | 0.5 |
| attr_d_beta1 | The beta1 in Adam for training the auxiliary discriminator | 0.5 |
| g_beta1 | The beta1 in Adam for training the generator | 0.5 |

Patients with SOFA scores below 5 are marked as SOFA^-^, while patients with SOFA scores above 8 are marked as SOFA^+^, and patients with SOFA scores in between are marked as SOFA°. The numbers of ICU stays marked SOFA^-^ and SOFA^+^ are 13,622 and 2,594, respectively.

To assess the utility of the synthetic data in a realistic clinical prediction setting, we consider a binary classification task where time series features are used to predict the SOFA score class. Only samples with either high (SOFA^+^) or low (SOFA^-^) SOFA scores are included, defining a binary classification task.

Classification was based on the MLP-LSTM-Static model described in Section 2.6. Test performance is measured using AUROC and AUPRC.

To select the hyperparameters of the MLP-LSTM-Static model, we conducted HO again using Optuna with tree-structured Parzen estimators, hyperband pruning, and a reduction factor of 2. We reserved 20% of the training data as a validation split for HO. The optimization was conducted until 200 trials were conducted, using the average precision of the classification of the validation set as the performance metric. Confidence intervals for the validation performance were obtained using bootstrapping (K=1,000).

## S4. Additional faithfulness metrics

Let *r*_*n,i,j*_ be the pairwise Pearson correlation coefficient of the *n*^th^ sample as defined in Equation 7, we define the mean normalized absolute difference (CorrMAD) and the mean normalized squared error (CorrMSE) as

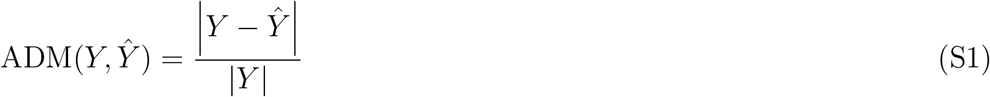

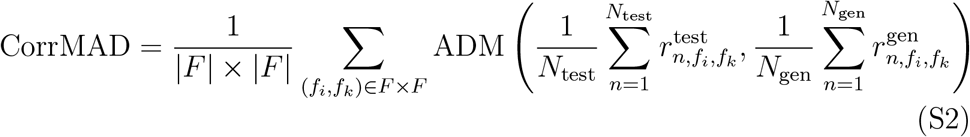

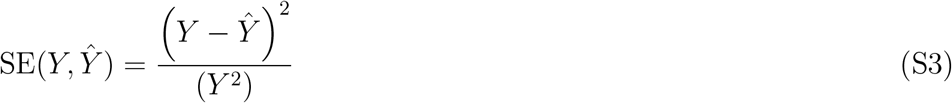

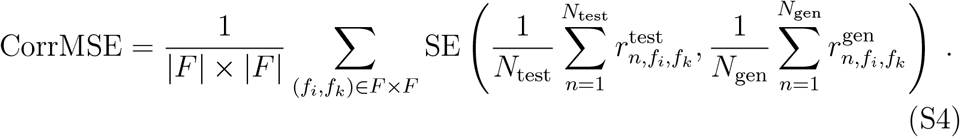

Analogous, we define the ACWSD metric for assessing the reconstruction of autocorrelation properties based on autocorrelation coefficients *r*_*n,f,h*_ as defined in Equation 12:

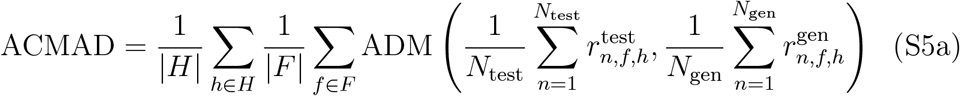

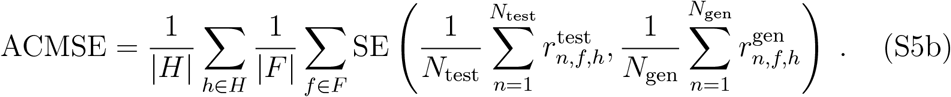

Corresponding metrics for assessing cross-correlation reconstruction based on coefficients 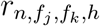 defined in Equation 15 are given as:

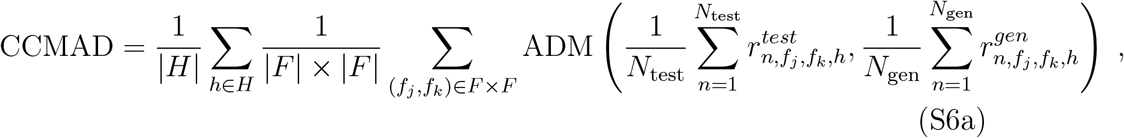

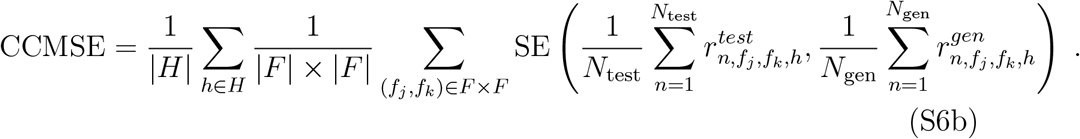

Lastly, we define corresponding metrics for static attributes as

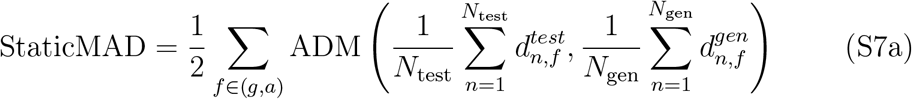

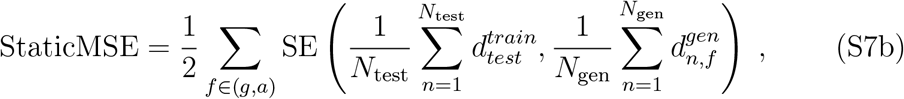

where *d*_*n,f*_ is an element of **d**_attr_ = (*d*_*i,f*_).

## S5. Additional results

**Table S5:** Performance of the MLP-LSTM-Static model in the downstream data utility task on the MIMIC-IV dataset. Performance was assessed using the area under precision-recall curve (AUPRC) and the area under receiver operator curve (AUROC). Performance was evaluated in two scenarios: *Train on Synthetic, Test on Real* : classifier was trained on synthetic data generated by the respective method and tested on real MIMIC-IV data. *Train on Real, Test on Synthetic*: classifier was trained on MIMIC-IV data and tested on the synthetic data generated by the respective method. The baseline *Train on Real, Test on Real* – classifier was trained and tested on MIMIC-IV data – is located on the first raw.

|  | AUROC |  | AUPRC |  |
| --- | --- | --- | --- | --- |
|  | Train on synth.,<br>test on real | Train on real,<br>test on synth. | Train on synth.,<br>test on real | Train on real,<br>test on synth. |
| Real | 0.727 ± 0.010 | 0.727 ± 0.010 | 0.922 ± 0.005 | 0.922 ± 0.005 |
| GHOSTS | **0.736 ± 0.010** | **0.699 ± 0.011** | **0.925 ± 0.005** | 0.907 ± 0.005 |
| GHOSTS-POST | 0.727 ± 0.010 | 0.674 ± 0.012 | 0.921 ± 0.005 | 0.897 ± 0.006 |
| DoppelGANger | 0.684 ± 0.011 | 0.671 ± 0.010 | 0.908 ± 0.006 | **0.912 ± 0.005** |
| HALO | 0.559 ± 0.012 | 0.574 ± 0.012 | 0.858 ± 0.007 | 0.867 ± 0.006 |

**Table S6:** Performance of the MLP-LSTM Static model in the data augmentation task on the MIMIC-IV dataset. Performance was assessed for different degrees of augmentation using the area under precision-recall curve (AUPRC) and the area under receiver operator curve (AUROC).

| Aug<br>frac | AUROC |  |  |  | Average Precision (AUPRC) |  |  |  |
| --- | --- | --- | --- | --- | --- | --- | --- | --- |
|  | GHOSTS | GHOSTS-POST | DoppelGANger | HALO | GHOSTS | GHOSTS-POST | DoppelGANger | HALO |
| 0.0 | 0.727 ± 0.010 | 0.727 ± 0.010 | 0.727 ± 0.010 | 0.727 ± 0.010 | 0.922 ± 0.005 | 0.922 ± 0.005 | 0.922 ± 0.005 | 0.922 ± 0.005 |
| 0.1 | **0.755 ± 0.010** | 0.563 ± 0.012 | 0.474 ± 0.012 | 0.744 ± 0.010 | **0.934 ± 0.004** | 0.852 ± 0.008 | 0.818 ± 0.008 | 0.931 ± 0.004 |
| 0.3 | **0.754 ± 0.010** | 0.566 ± 0.012 | 0.471 ± 0.013 | 0.742 ± 0.010 | **0.934 ± 0.004** | 0.853 ± 0.008 | 0.816 ± 0.008 | 0.930 ± 0.004 |
| 0.5 | **0.753 ± 0.010** | 0.567 ± 0.012 | 0.469 ± 0.012 | 0.741 ± 0.010 | **0.934 ± 0.004** | 0.854 ± 0.008 | 0.815 ± 0.009 | 0.930 ± 0.005 |
| 0.8 | **0.754 ± 0.010** | 0.569 ± 0.012 | 0.468 ± 0.012 | 0.739 ± 0.010 | **0.934 ± 0.004** | 0.855 ± 0.008 | 0.815 ± 0.009 | 0.929 ± 0.005 |
| 1.0 | **0.754 ± 0.010** | 0.569 ± 0.012 | 0.468 ± 0.012 | 0.740 ± 0.010 | **0.934 ± 0.004** | 0.855 ± 0.008 | 0.814 ± 0.008 | 0.929 ± 0.004 |
| 2.0 | **0.754 ± 0.010** | 0.569 ± 0.013 | 0.468 ± 0.013 | 0.739 ± 0.010 | **0.934 ± 0.004** | 0.855 ± 0.008 | 0.814 ± 0.008 | 0.929 ± 0.004 |
| 5.0 | **0.753 ± 0.009** | 0.569 ± 0.013 | 0.468 ± 0.012 | 0.739 ± 0.010 | **0.934 ± 0.004** | 0.855 ± 0.008 | 0.814 ± 0.008 | 0.929 ± 0.004 |
| 10.0 | **0.753 ± 0.010** | 0.568 ± 0.012 | 0.468 ± 0.012 | 0.739 ± 0.010 | **0.934 ± 0.004** | 0.855 ± 0.008 | 0.815 ± 0.008 | 0.929 ± 0.004 |

**Table S7:**
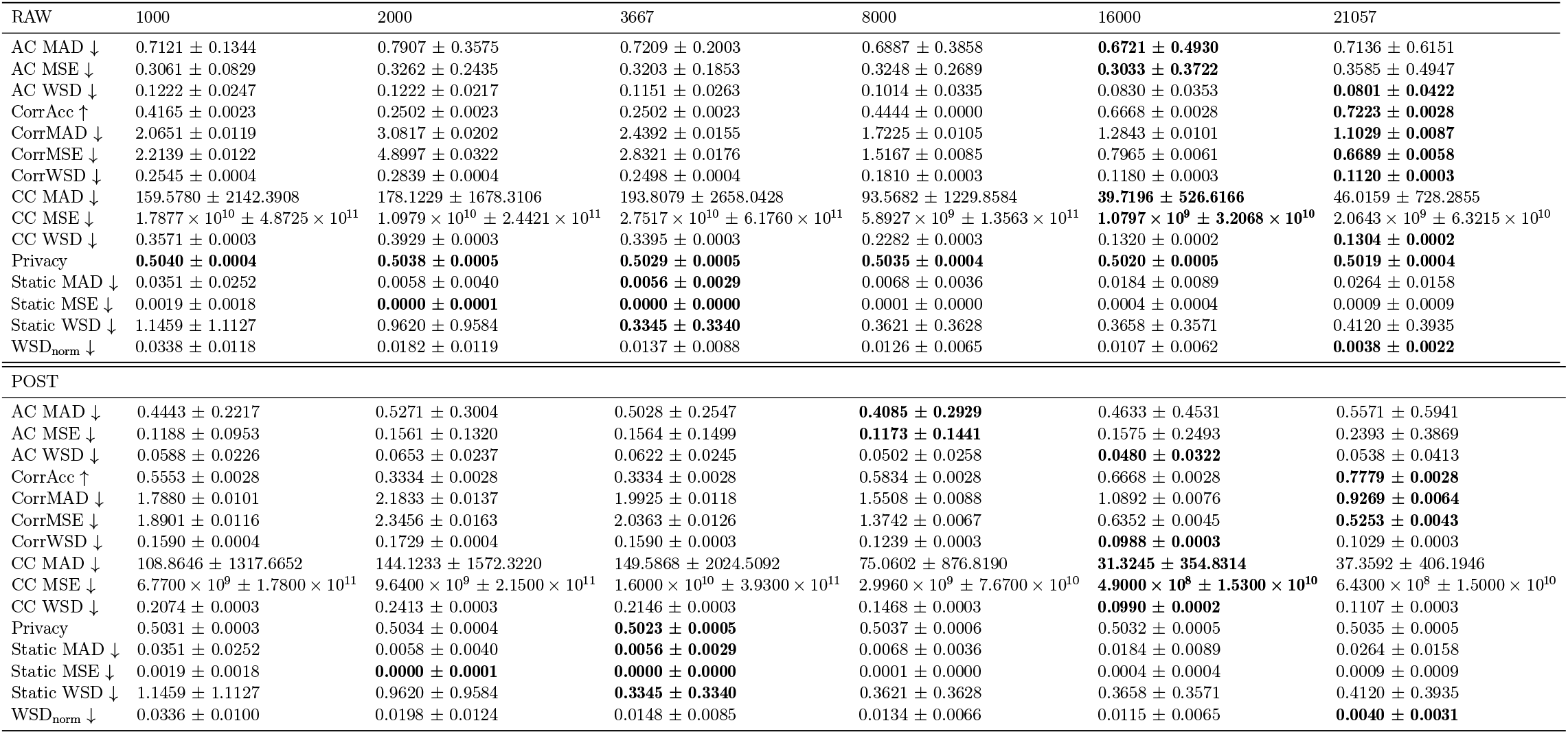
Comparison of different training set sizes in combiantion with the GHOST_RAW and GHOST_POST models in terms of distributional faithfulness on the MIMIC-IV dataset. Both models were trained on 1000, 2000, 3667, 8000, 16000 and 21057 ICU stays.

**Table S8:**
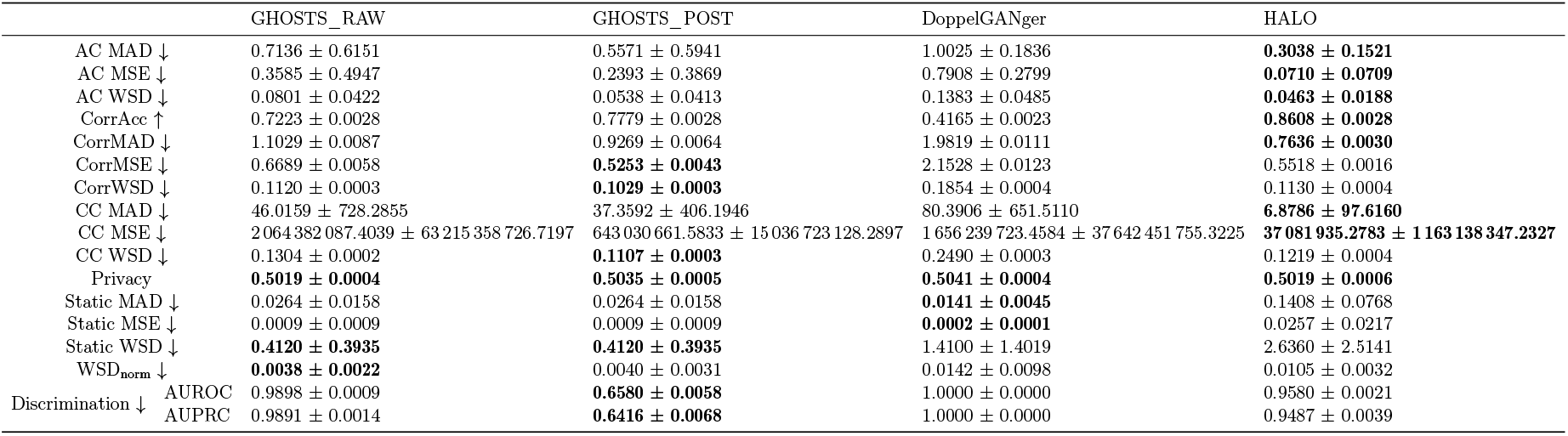
Quantitative evaluation of how faithfully synthetic MIMIC-IV data generated by DoppelGANger, GHOSTS without postprocessing (GHOSTS_RAW) and GHOSTS with postprocessing (GHOSTS_POST) reproduce the distribution of real MIMIC-IV data. WSD_norm_ refers to the normalized Wasserstein distance between univariate distributions of raw values of individual time series features presented in Equation (5). CorrAcc, CorrMSE, CorrMAD, and CorrWSD refer to correlation accuracy, mean squared error, mean absolute difference, and Wasserstein distance between distributions of real and synthetic |*F*| ×|*F*| temporal Pearson correlations between features, corresponding to Equations (10), (S4), (S2), and (11). Similarly, AC_{MAD,MSE,WSD}, CC_{MAD,MSE,WSD} denote auto- and correlation similarities derived from |*F*| ×|*F*| × *N*_lag_ lagged cross-covariance matrices, respectively, corresponding to Equations (14), (S5), (16) and (S6). Privacy refers to the Equation (20). Static_{MAD,MSE,WSD} denote comparisons between univariate distributions of static attributes presented in Equations (17) and (S7). Data are reported as mean ± standard error of the mean, and ↓/↑ indicate that lower/higher values are better, respectively. Uncertainty was estimated using the bootstrap method (K=1,000)

**Table S9:** Quantitative evaluation of how faithfully synthetic EICU data generated by HALO, DoppelGANger, GHOSTS without postprocessing (GHOSTS_RAW) and GHOSTS with postprocessing (GHOSTS_POST) reproduce the distribution of real EICU data. WSD_norm_ refers to the normalized Wasserstein distance between univariate distributions of raw values of individual time series features presented in Equation (5). CorrAcc, CorrMSE, CorrMAD, and CorrWSD refer to correlation accuracy, mean squared error, mean absolute difference, and Wasserstein distance between distributions of real and synthetic |*F*| × |*F*| temporal Pearson correlations between features, corresponding to Equations (10), (S4), (S2), and (11). Similarly, AC_{MAD,MSE,WSD}, CC_{MAD,MSE,WSD} denote auto- and correlation similarity derived from |*F*| × |*F*| × *N*_lag_ lagged cross-covariance matrices, respectively, corresponding to Equations (14), (S5), (16) and (S6). Privacy refers to the Equation (20). Static_{MAD,MSE,WSD} denote comparisons between univariate distributions of static attributes presented in Equations (17) and (S7). Discrimination AUROC and AUPRC refers to discriminaboloty as measured by area under receiver operator curve and area under precision-recall curve, described in Equations (18) and (19), respectively. Data are reported as mean ± standard error of the mean, and ↓/↑ indicate that lower/higher values are better, respectively. Uncertainty was estimated using the bootstrap method (K=1,000)

|  | GHOSTS_RAW | GHOSTS_POST | DoppelGANger | HALO |
| --- | --- | --- | --- | --- |
| AC MAD ↓ | 0.5413 ± 0.1677 | **0.3709 ± 0.1926** | 0.6307 ± 0.2545 | 0.4073 ± 0.2104 |
| AC MSE ↓ | 0.2130 ± 0.1293 | **0.1108 ± 0.1189** | 0.4133 ± 0.3275 | 0.1803 ± 0.1645 |
| AC WSD ↓ | 0.0952 ± 0.0243 | **0.0603 ± 0.0256** | 0.1103 ± 0.0482 | 0.0727 ± 0.0417 |
| CorrAcc ↑ | 0.4762 ± 0.0162 | 0.5595 ± 0.0162 | 0.4985 ± 0.0108 | **0.7512 ± 0.0159** |
| CorrMAD ↓ | 1.6712 ± 0.0334 | 1.4927 ± 0.0237 | 1.7451 ± 0.0267 | **0.8311 ± 0.0049** |
| CorrMSE ↓ | 1.6616 ± 0.0405 | 1.3946 ± 0.0243 | 1.8682 ± 0.0284 | **0.6169 ± 0.0039** |
| CorrWSD ↓ | 0.2138 ± 0.0010 | **0.1622 ± 0.0010** | 0.2107 ± 0.0010 | 0.1826 ± 0.0012 |
| CC MAD ↓ | 33.4142 ± 35.8454 | 34.3638 ± 112.6498 | 28.2234 ± 76.9366 | **1.7245 ± 1.5921** |
| CC MSE ↓ | 5 196 721.0877 ± 56 978 102.5071 | 49 753 341.0963 ± 1 447 582 100.4845 | 23 318 407.4318 ± 643 032 299.1374 | **9867.7328 ± 118 751.0474** |
| CC WSD ↓ | 0.3007 ± 0.0008 | 0.2288 ± 0.0007 | 0.2460 ± 0.0009 | **0.2021 ± 0.0011** |
| Privacy | 0.5047 ± 0.0014 | 0.5048 ± 0.0013 | 0.5053 ± 0.0012 | 0.5081 ± 0.0016 |
| Static MAD ↓ | 0.0099 ± 0.0085 | 0.0099 ± 0.0085 | **0.0093 ± 0.0058** | 0.1616 ± 0.0599 |
| Static MSE ↓ | 0.0002 ± 0.0003 | 0.0002 ± 0.0003 | **0.0001 ± 0.0002** | 0.0297 ± 0.0195 |
| Static WSD ↓ | **0.3906 ± 0.3883** | **0.3906 ± 0.3883** | 0.6537 ± 0.6514 | 3.4663 ± 3.3315 |
| $WSD_{\text{norm}}$ ↓ | **0.0067 ± 0.0042** | **0.0067 ± 0.0045** | 0.0220 ± 0.0127 | 0.0159 ± 0.0154 |
| Discrimination ↓ AUROC | 0.7699 ± 0.0119 | **0.5688 ± 0.0146** | 0.9328 ± 0.0066 | 0.7148 ± 0.0134 |
| AUPRC | 0.7559 ± 0.0174 | **0.5944 ± 0.0192** | 0.8967 ± 0.0147 | 0.6489 ± 0.0191 |

## S6. Distribution comparisons

**Figure S1:**
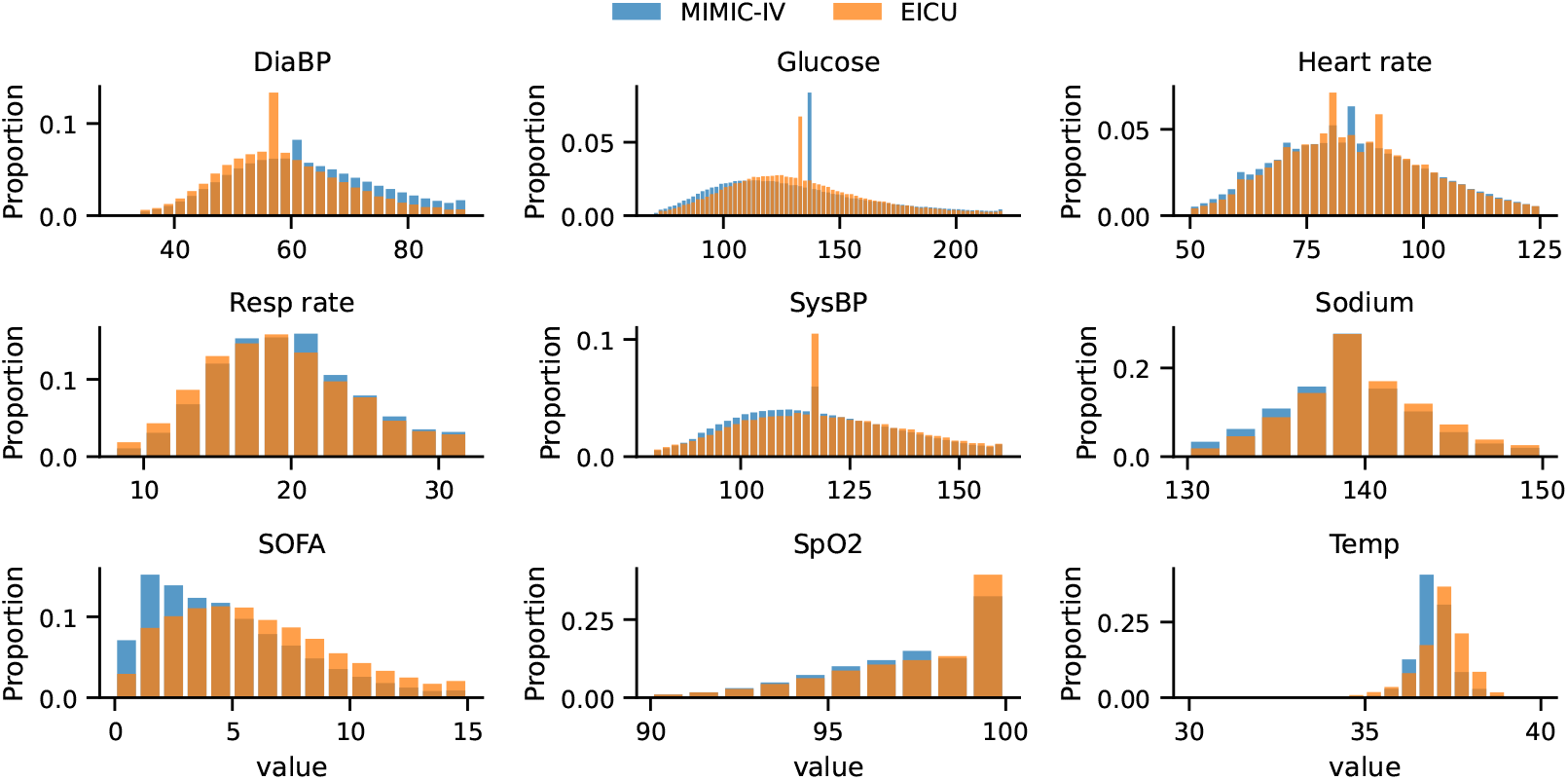
Comparison of the distributions of 9 clinical variables between the real MIMIC-IV and eICU datasets. The data points are filtered between 1 and 99 quantiles for clarity. The aggregated statistics are normalized such that such that bar heights sum to 1. Abbreviations: DiaBP - diastolic blood pressure, Resp rate - respiratory rate, SOFA - Sequential Organ Failure Assessment (SOFA) score, SpO2 - oxygen saturation, SysBP - systolic blood pressure, Temp - temperature.

**Figure S2:**
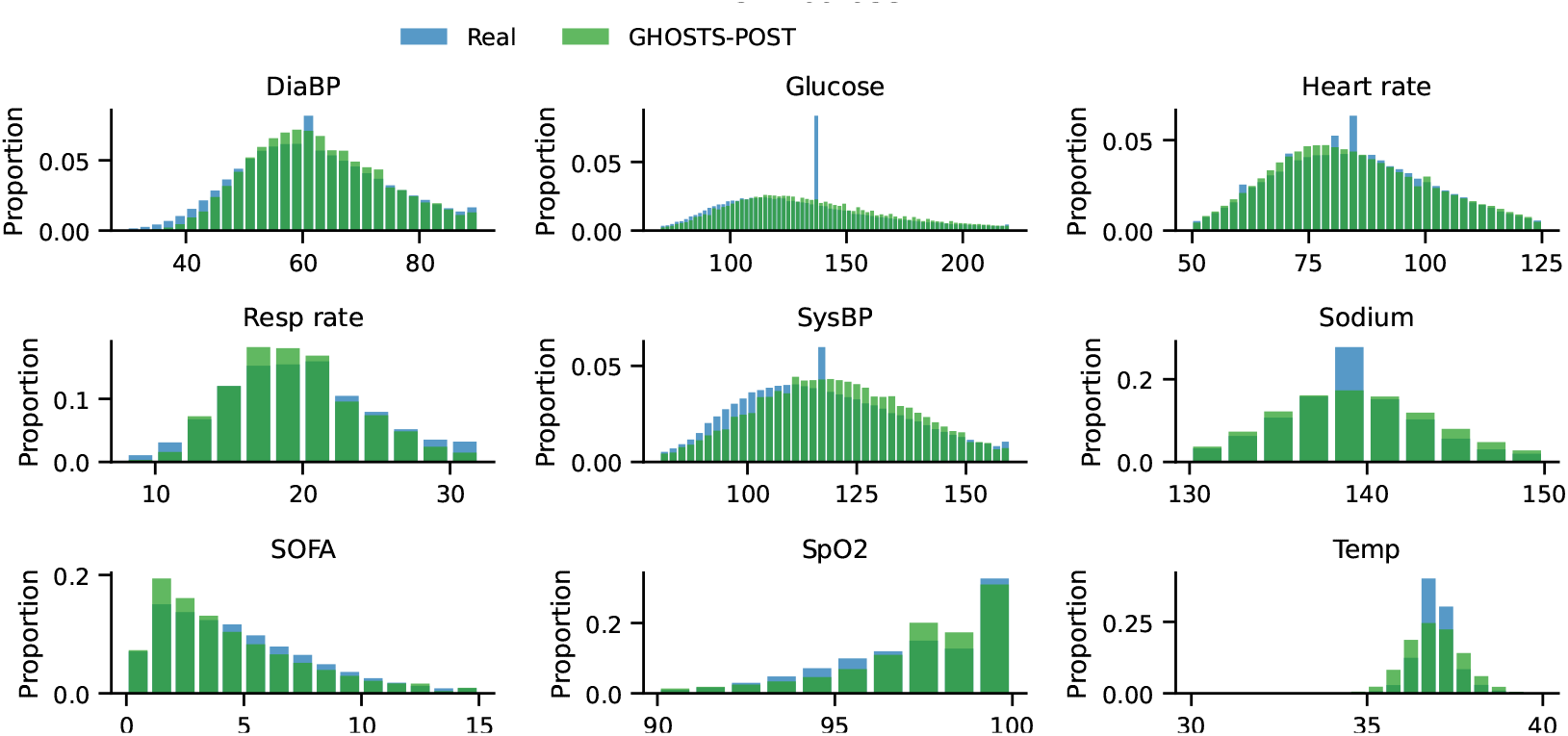
Comparison of the distributions of 9 clinical variables between the real MIMIC-IV data and synthetic GHOSTS-POST data. The data points are filtered between 1 and 99 quantiles for clarity. The aggregated statistics are normalized such that such that bar heights sum to 1. Abbreviations: DiaBP - diastolic blood pressure, Resp rate - respiratory rate, SOFA - Sequential Organ Failure Assessment (SOFA) score, SpO2 - oxygen saturation, SysBP - systolic blood pressure, Temp - temperature.

**Figure S3:**
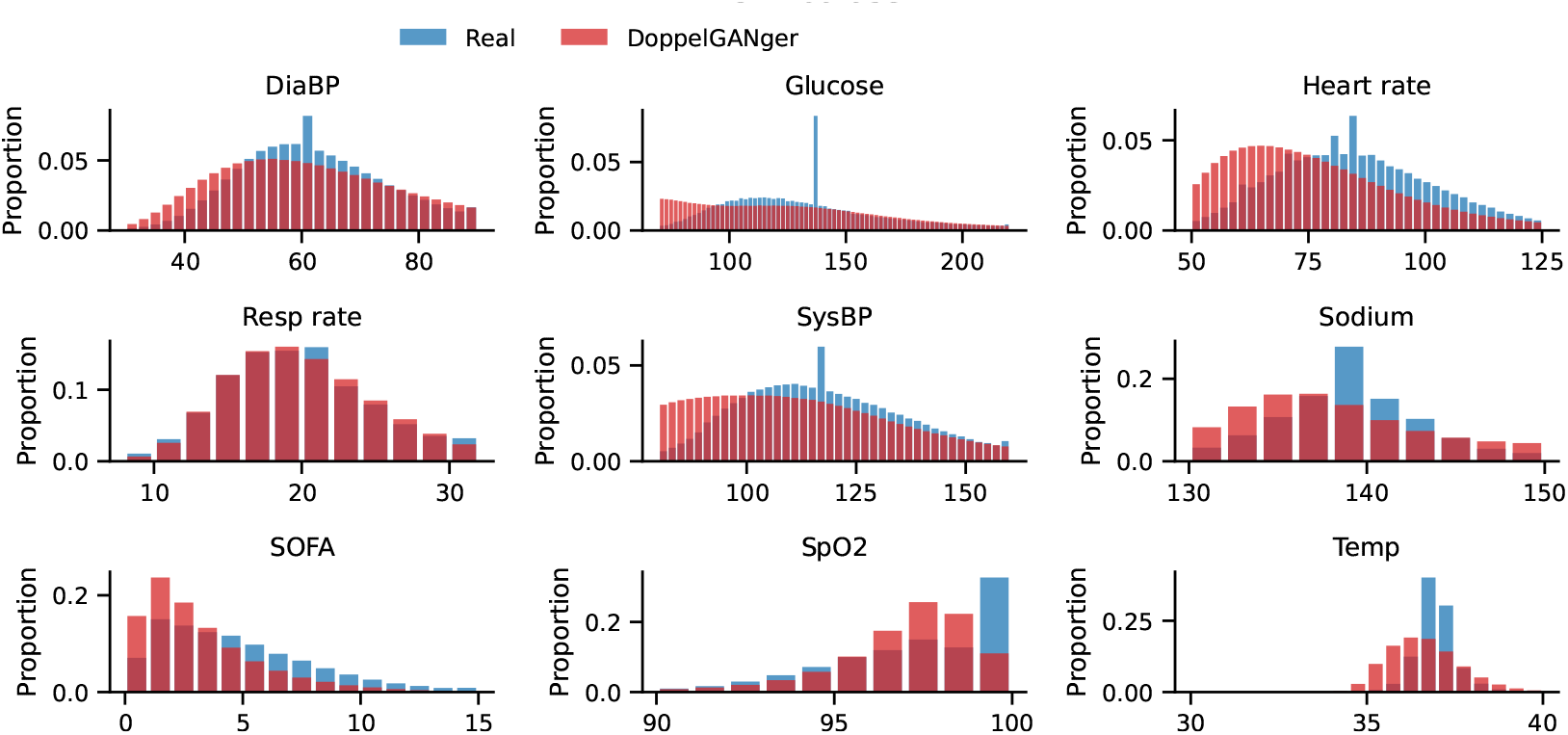
Comparison of the distributions of 9 clinical variables between the real MIMIC-IV data and synthetic DoppelGANger data. The data points are filtered between 1 and 99 quantiles for clarity. The aggregated statistics are normalized such that such that bar heights sum to 1. Abbreviations: DiaBP - diastolic blood pressure, Resp rate - respiratory rate, SOFA - Sequential Organ Failure Assessment (SOFA) score, SpO2 - oxygen saturation, SysBP - systolic blood pressure, Temp - temperature.

**Figure S4:**
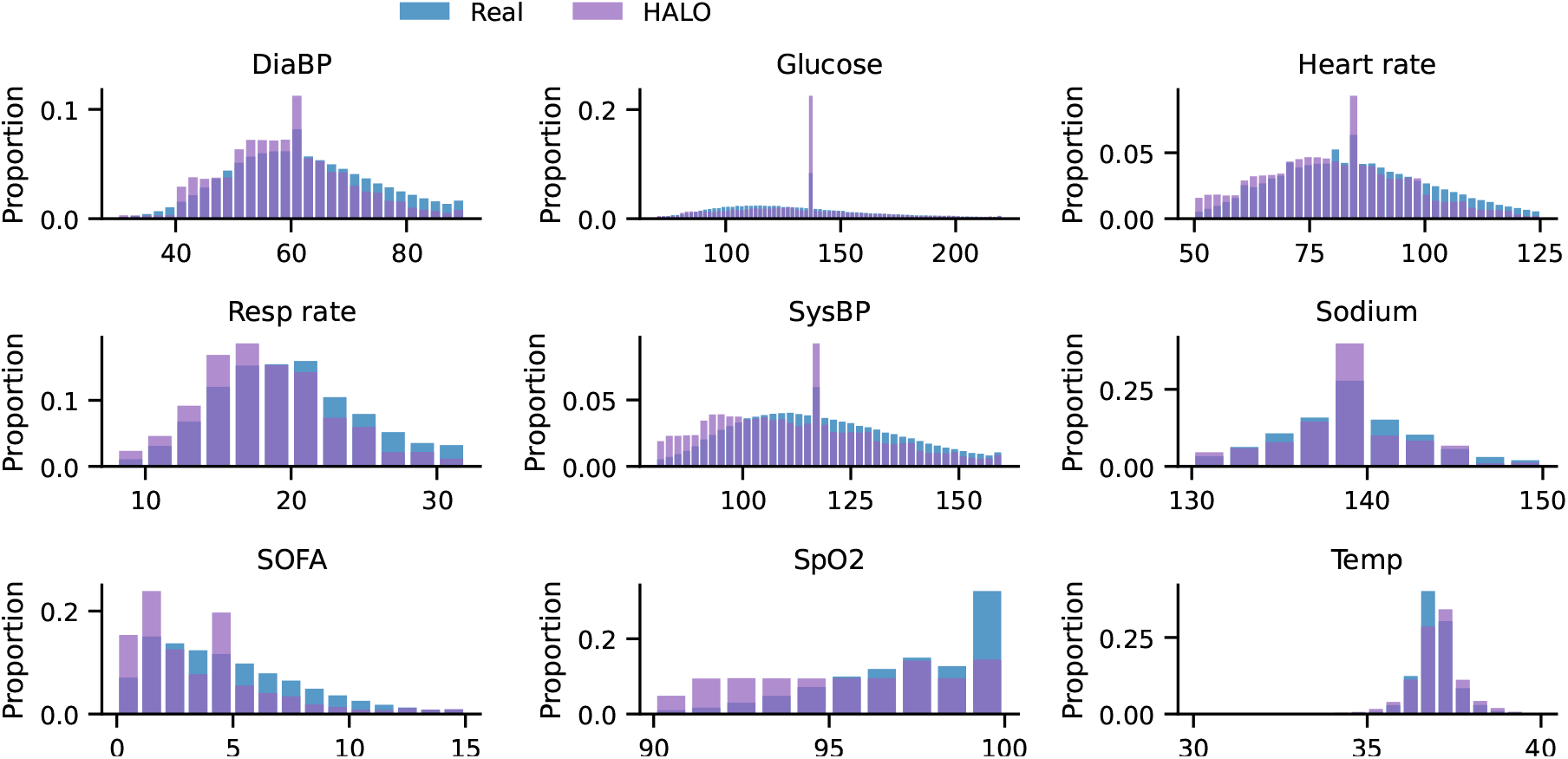
Comparison of the distributions of 9 clinical variables between the real MIMIC-IV data and synthetic HALO data. The data points are filtered between 1 and 99 quantiles for clarity. The aggregated statistics are normalized such that such that bar heights sum to 1. Abbreviations: DiaBP - diastolic blood pressure, Resp rate - respiratory rate, SOFA - Sequential Organ Failure Assessment (SOFA) score, SpO2 - oxygen saturation, SysBP - systolic blood pressure, Temp - temperature.

**Figure S5:**
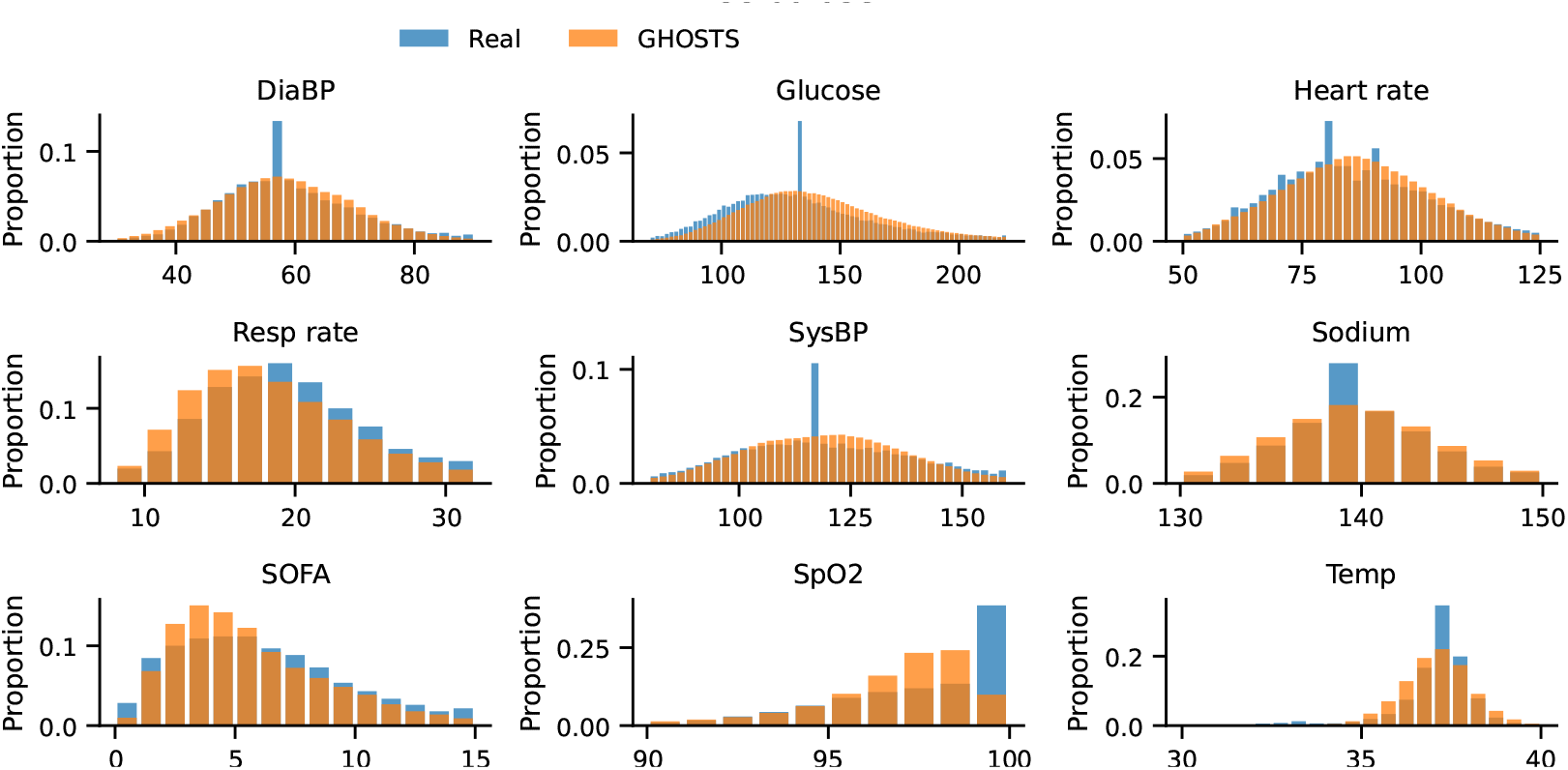
Comparison of the distributions of 9 clinical variables between the real eICU data and synthetic GHOSTS data. The data points are filtered between 1 and 99 quantiles for clarity. The aggregated statistics are normalized such that such that bar heights sum to 1. Abbreviations: DiaBP - diastolic blood pressure, Resp rate - respiratory rate, SOFA - Sequential Organ Failure Assessment (SOFA) score, SpO2 - oxygen saturation, SysBP - systolic blood pressure, Temp - temperature.

**Figure S6:**
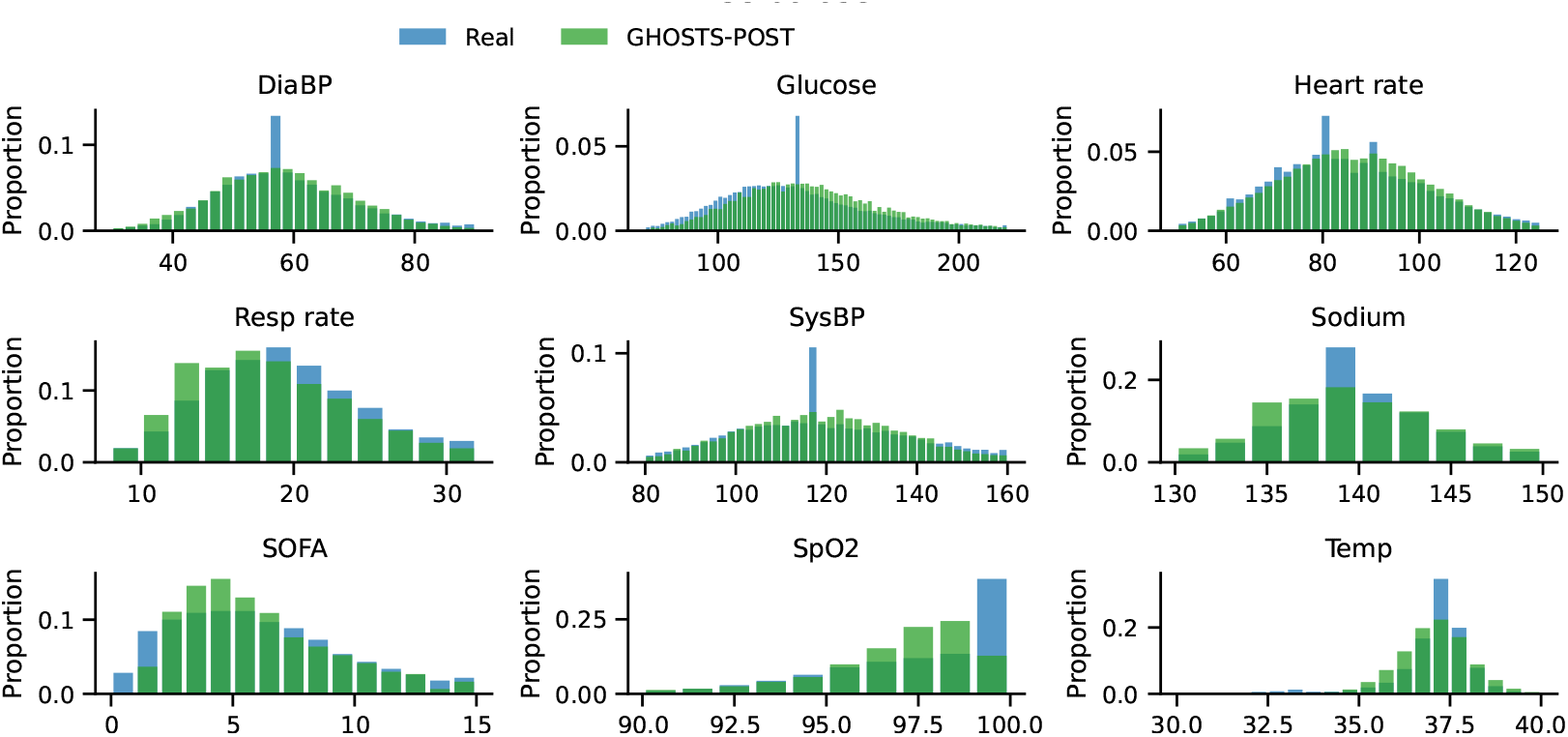
Comparison of the distributions of 9 clinical variables between the real eICU data and synthetic GHOSTS-POST data. The data points are filtered between 1 and 99 quantiles for clarity. The aggregated statistics are normalized such that such that bar heights sum to 1. Abbreviations: DiaBP - diastolic blood pressure, Resp rate - respiratory rate, SOFA - Sequential Organ Failure Assessment (SOFA) score, SpO2 - oxygen saturation, SysBP - systolic blood pressure, Temp - temperature.

**Figure S7:**
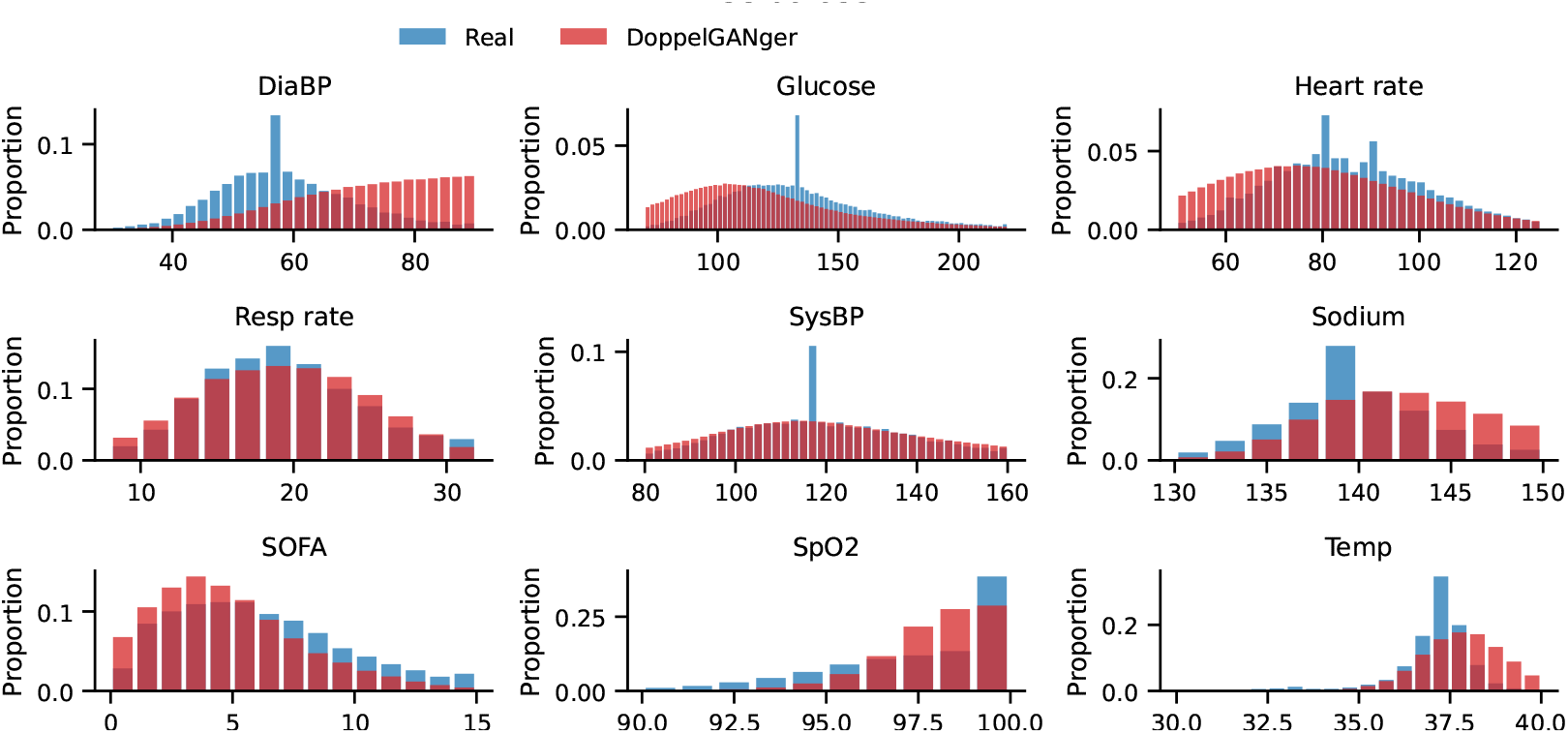
Comparison of the distributions of 9 clinical variables between the real eICU data and synthetic DoppelGANger data. The data points are filtered between 1 and 99 quantiles for clarity. The aggregated statistics are normalized such that such that bar heights sum to 1. Abbreviations: DiaBP - diastolic blood pressure, Resp rate - respiratory rate, SOFA - Sequential Organ Failure Assessment (SOFA) score, SpO2 - oxygen saturation, SysBP - systolic blood pressure, Temp - temperature.

**Figure S8:**
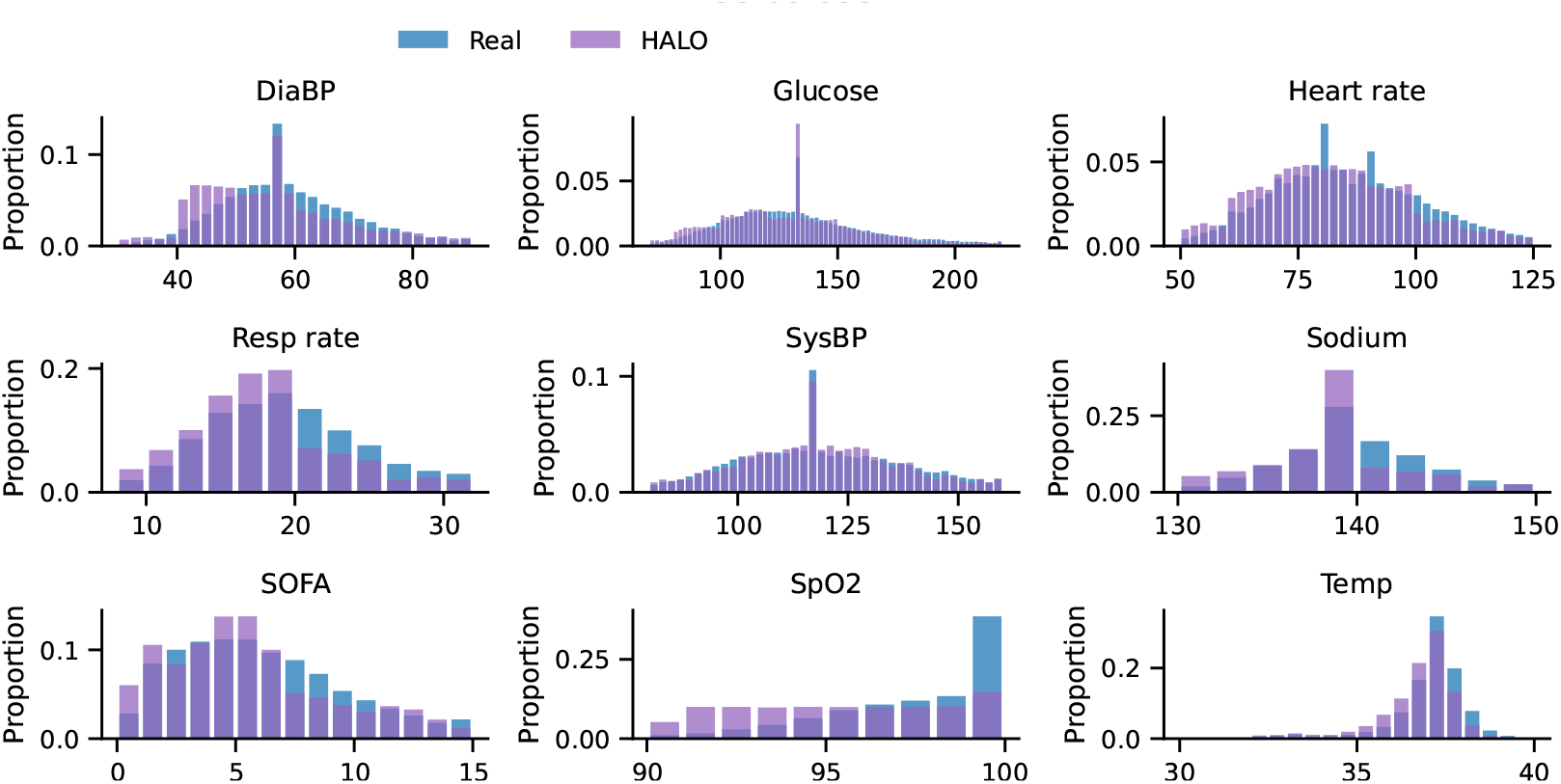
Comparison of the distributions of 9 clinical variables between the real eICU data and synthetic HALO data. The data points are filtered between 1 and 99 quantiles for clarity. The aggregated statistics are normalized such that such that bar heights sum to 1. Abbreviations: DiaBP - diastolic blood pressure, Resp rate - respiratory rate, SOFA - Sequential Organ Failure Assessment (SOFA) score, SpO2 - oxygen saturation, SysBP - systolic blood pressure, Temp - temperature.

